# BugSigDB captures patterns of differential abundance across a broad range of host-associated microbial signatures

**DOI:** 10.1101/2022.10.24.22281483

**Authors:** Ludwig Geistlinger, Chloe Mirzayi, Fatima Zohra, Rimsha Azhar, Shaimaa Elsafoury, Claire Grieve, Jennifer Wokaty, Samuel David Gamboa-Tuz, Pratyay Sengupta, Isaac Hecht, Aarthi Ravikrishnan, Rafael Gonçalves, Eric Franzosa, Karthik Raman, Vincent Carey, Jennifer B. Dowd, Heidi E. Jones, Sean Davis, Nicola Segata, Curtis Huttenhower, Levi Waldron

**Author notes:** Corresponding author: Levi Waldron.

## Abstract

The literature of human and other host-associated microbiome studies is expanding rapidly, but systematic comparisons among published results of host-associated microbiome signatures of differential abundance remain difficult. We present BugSigDB, a community-editable database of manually curated microbial signatures from published differential abundance studies, accompanied by information on study geography, health outcomes, host body site, and experimental, epidemiological, and statistical methods using controlled vocabulary. The initial release of the database contains >2,500 manually curated signatures from >600 published studies on three host species, enabling high-throughput analysis of signature similarity, taxon enrichment, co-occurrence and co-exclusion, and consensus signatures. These data allow assessment of microbiome differential abundance within and across experimental conditions, environments, or body sites. Database-wide analysis reveals experimental conditions with the highest level of consistency in signatures reported by independent studies and identifies commonalities among disease-associated signatures including frequent introgression of oral pathobionts into the gut.

## Main

Despite substantial progress in experimental techniques and computational methods for culture-independent profiling of the human microbiome, the analysis and interpretation of microbial differential abundance studies remains challenging. A large body of experimental and observational studies on humans and in animal models has reported associations between host-associated microbiomes and the onset, progression, and treatment of a variety of diseases including atherosclerosis^1^, cardiovascular diseases^2^, cancers^3^, and diabetes^4^. This growing body of published results provides opportunities for synthesis of accumulated knowledge, identification of common patterns across different diseases and exposures, and interpretation of new studies by comparison to previous results. However, without a systematic catalog of published differential abundance results, even identical microbial signatures reported in different research fields are unlikely to be noticed. Even within research fields, systematic review in the absence of a catalog or common reporting of differential abundance results are time-consuming, static, and generally do not summarize all taxa reported.

This situation has parallels to early challenges in the interpretation of differential gene expression analysis^5^, which have been addressed in the field by Gene Set Enrichment Analysis (GSEA). GSEA allows the comparison of coherent expression patterns among predefined gene signatures that share a biological function, property, or that were identified together by a previous study^6,7^. GSEA is a key tool in gene expression data analysis^6^, with a wide range of subsequent methods^8^ to account for correlations between genes^9^, redundancy of functional annotation^10^, different types of null hypothesis^11^, and the application of GSEA for the analysis of genomic regions^12^, metabolomic data^13^, and disease phenotypes^14^.

Analogously, differential microbial abundance analysis can yield lists or “signatures” of microbial clades at multiple taxonomic levels that are associated with a phenotype of interest. The properties shared by these clades are often not obvious, but could include common environmental exposures, metabolic or ecological requirements, or physiological characteristics. Although nascent attempts to apply concepts of GSEA to results of microbiome differential abundance analysis exist^15–18^, major obstacles have prevented their broad utility and adoption. The most significant obstacle has been the lack of comprehensive databases of signatures designed for enrichment analysis, such as those available for GSEA including GO^19^, KEGG^20^, MSigDB^21,22^, and GeneSigDB^23^. Several databases provide significant information on microbial physiology and morphology^24–27^, but are not designed for enrichment analysis and, by design, exclude the vast majority of experimentally-derived microbial signatures associated with cancer, inflammation, diet, or other conditions studied in human and other host-associated microbiome research.

This study provides two main contributions to enable high-throughput comparison of published microbial signatures. First, it describes BugSigDB, a database of published microbial signatures of sufficient scale and diversity to capture replicable patterns of differential abundance across a broad spectrum of the host-associated microbiome literature. BugSigDB provides curated published signatures of differentially abundant microbes associated with a wide range of health outcomes, pharmaceutical usage (e.g. antibiotics), experiments on animal models, randomized clinical trials, and microbial attributes, and is built on the technology of Wikipedia to allow community contributions, revisions, and review for quality control. Second, we provide a systematic analysis of the results reported by hundreds of published microbiome studies, identifying replicated patterns even across 16S amplicon and shotgun sequencing approaches, demonstrating that interpretation of new microbiome studies can be supported by systematic comparison to previously published signatures. Database-wide analysis revealed common patterns of microbe co-occurrence and mutual exclusivity within signatures, and identified antibiotic treatment and HIV infection as the experimental conditions with the highest level of consistency in signatures reported by independent studies. “Bug set” enrichment analysis of 10 individual-participant colorectal cancer-associated fecal microbiome datasets (*N* = 663) detected published meta-analysis signatures used as positive controls, supporting direct application of methods adapted from gene set analysis as well as taxonomy-aware enrichment methods. This analysis also identified commonalities among signatures of other diseases, including elevated frequency of oral pathobionts in the gut, and identified the most common patterns of co-occurrence and mutual exclusion across all conditions, related conditions, or uniquely to one condition. Together, the BugSigDB database and analysis methods described here improve the interpretation of new microbiome studies by systematic comparison to published microbial signatures.

## Results

### A curated database of published microbial signatures

BugSigDB comprises a comprehensive database of manually-curated, host-associated microbial signatures from published microbiome studies, currently of human, mouse, and rat (Figure 1). The database has been simultaneously developed and expanded over the course of 4+ years with contributions from more than 25 curators trained in-house, currently containing >2,500 microbial signatures extracted from >600 scientific articles (Figure 1A). The curated papers cover two decades of microbiome research, with the majority of studies being published in the last 5 years (459 / 628 articles, 73.1%, Supplementary Figure S1). Among them are microbiome studies of participants from more than 50 different countries, with more than 50% of the studies originating from China and the USA (201 and 157 studies, respectively; Figure 1B).

**Figure 1:**
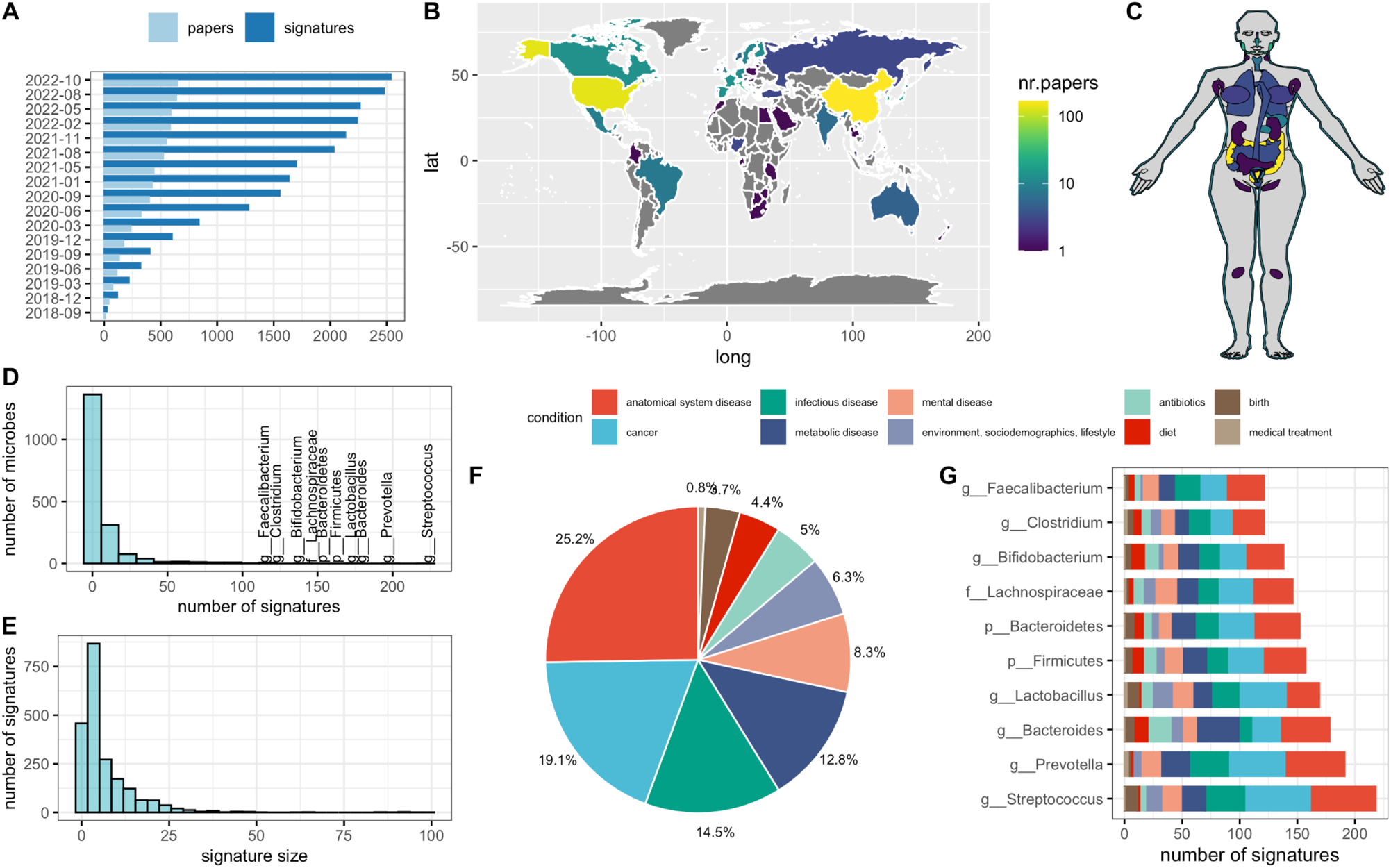
BugSigDB – a curated database of experimentally-derived microbial signatures. **(A)** BugSigDB is a community-editable collection of published microbiome studies reporting differentially abundant host-associated microbiota (including bacteria, archaea, fungi, protists, and viruses) that currently includes >2,500 microbial signatures extracted from >600 scientific articles over the course of 4+ years. These papers report microbiome studies of **(B)** participants representing different countries and ethnogeographies, and **(C)** microbiome samples from different human body sites. **(D)** Number of signatures associated with a specific microbe, with the top 10 most frequently reported microbes annotated. BugSigDB signatures contain taxonomic levels from phylum to strain, standardized based on the NCBI Taxonomy^29^. **(E)** Signature sizes, with more than 50% of the signatures containing 5 or more microbes. **(F)** Percentage of signatures annotated to major disease categories when classifying the study condition associated with each signature according to the Experimental Factor Ontology^32^. **(G)** The top 10 most frequently reported microbes and the number of associated signatures, stratified by disease category.

Studying microbiome samples from 14 broad body areas comprising more than 60 refined anatomical sites according to the UBERON Anatomy Ontology^28^ (Supplementary Table S1), the majority of studies in BugSigDB analyzed gut (440 / 628 studies, 70.1%), oral (80 studies, 12.8%), and vaginal microbiome samples (59 studies, 9.4%, Figure 1C). The signatures are generated by both 16S amplicon sequencing (92.5%) and metagenomic shotgun sequencing (MGX, 7.5%, Table 1), and contain taxonomic levels from phylum to strain standardized based on the NCBI Taxonomy^29^ (Supplementary Figure S2). BugSigDB is implemented as a semantic MediaWiki^30^ web interface available at https://bugsigdb.org, supporting data entry, semantic validation, quality control, and web-based programmatic access to annotations for studies, experiments, signatures, and individual taxa (Supplementary Figure S3 and Methods, Section *Data entry, validation, and access*).

**Table 1.**
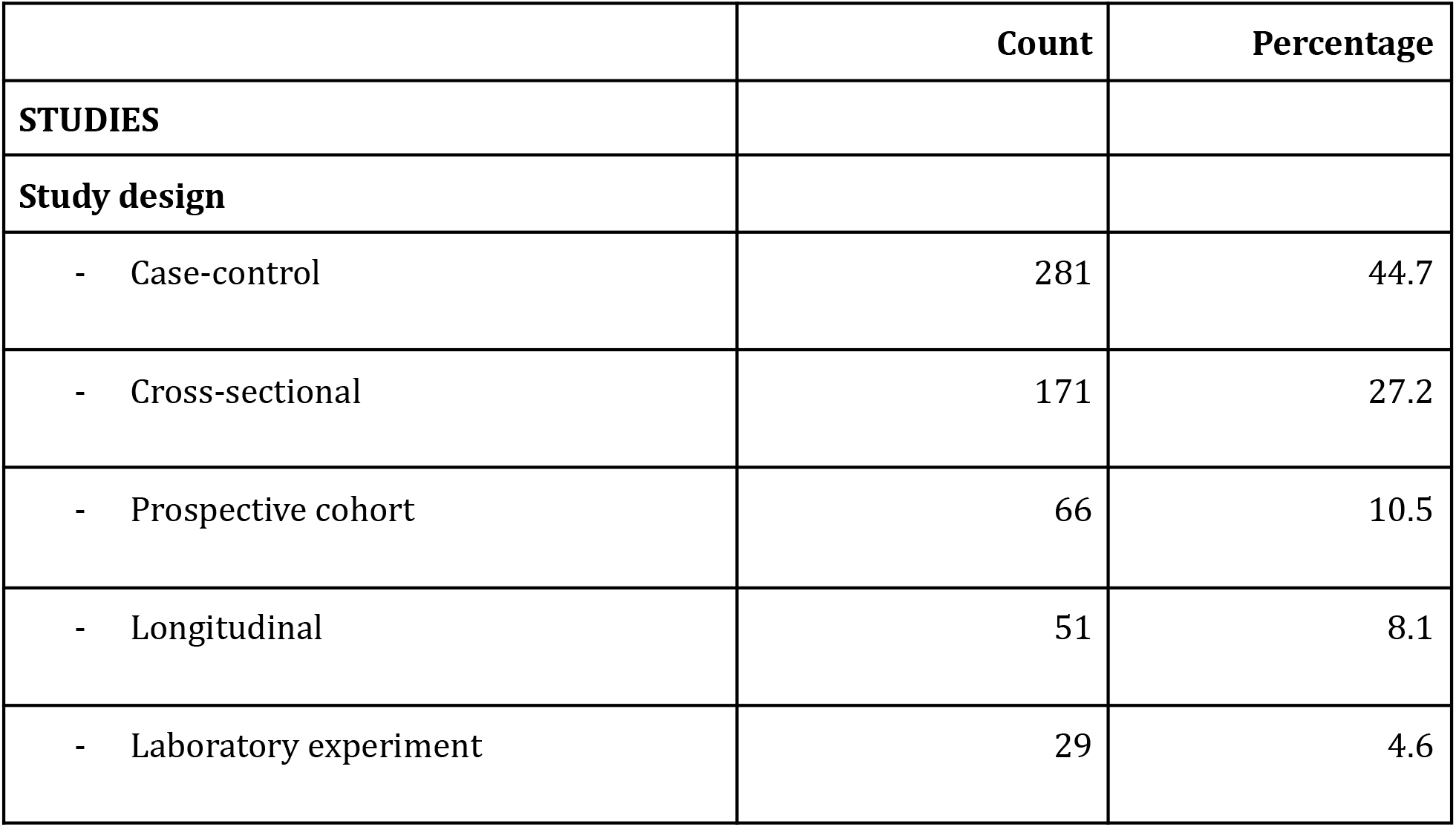
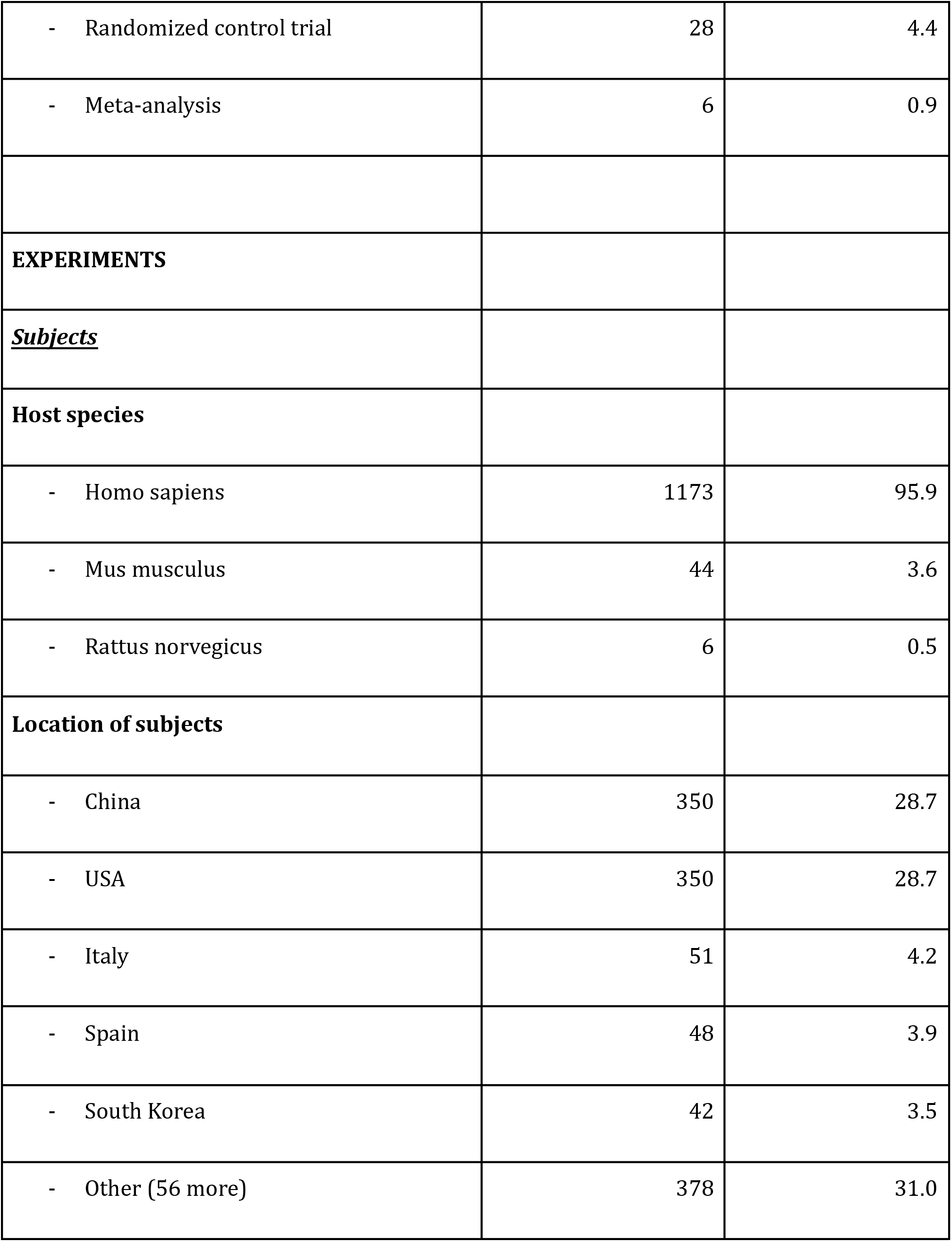
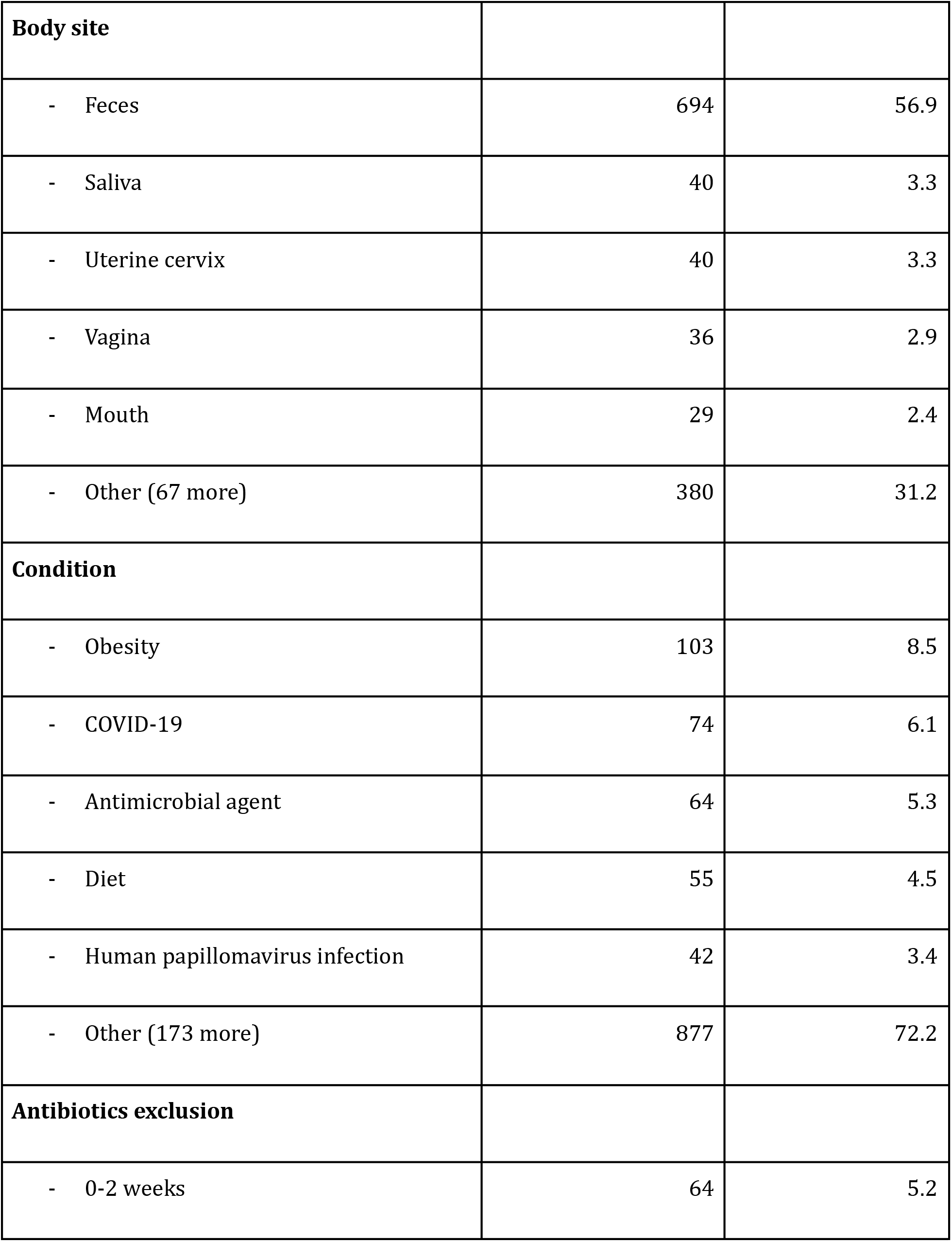
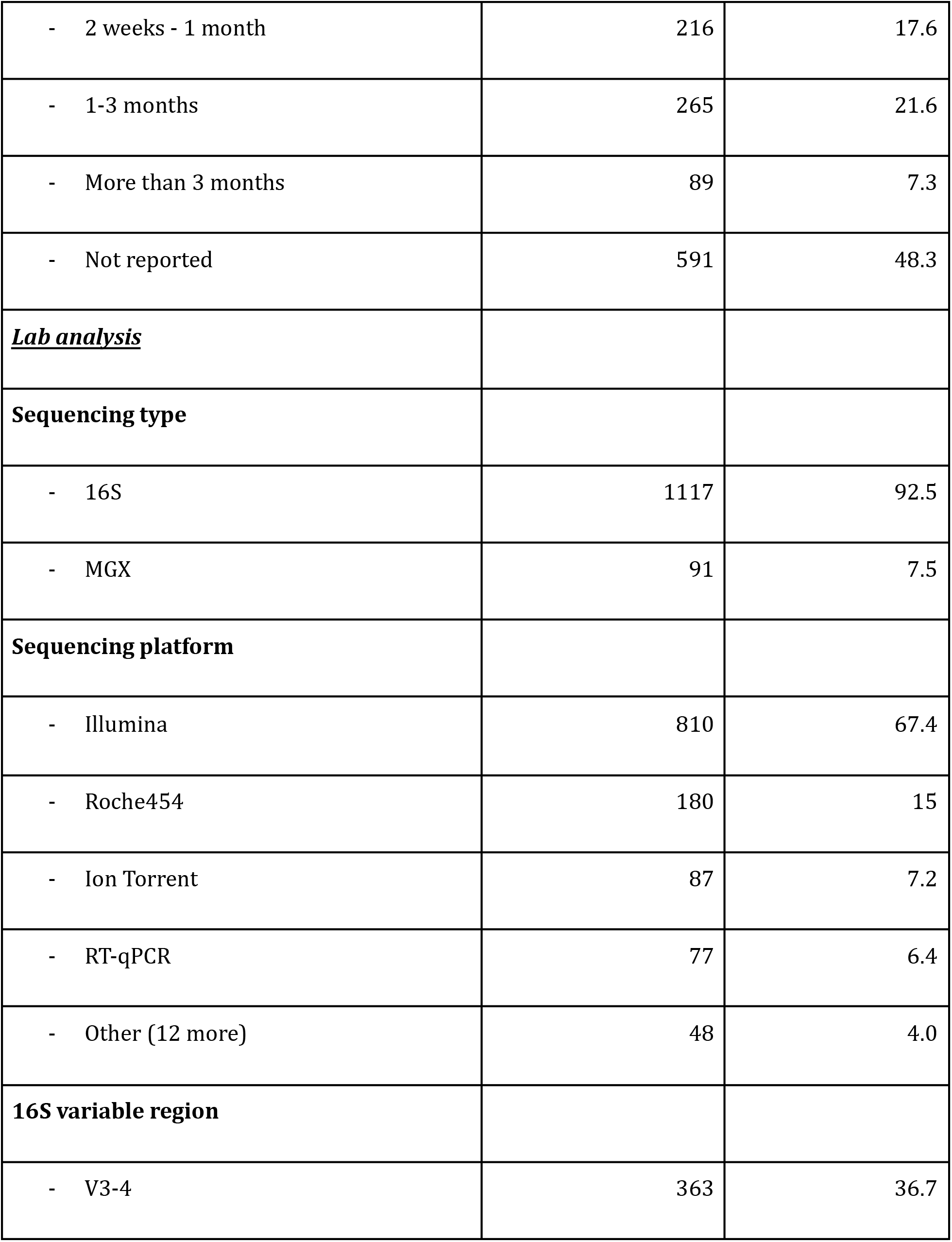
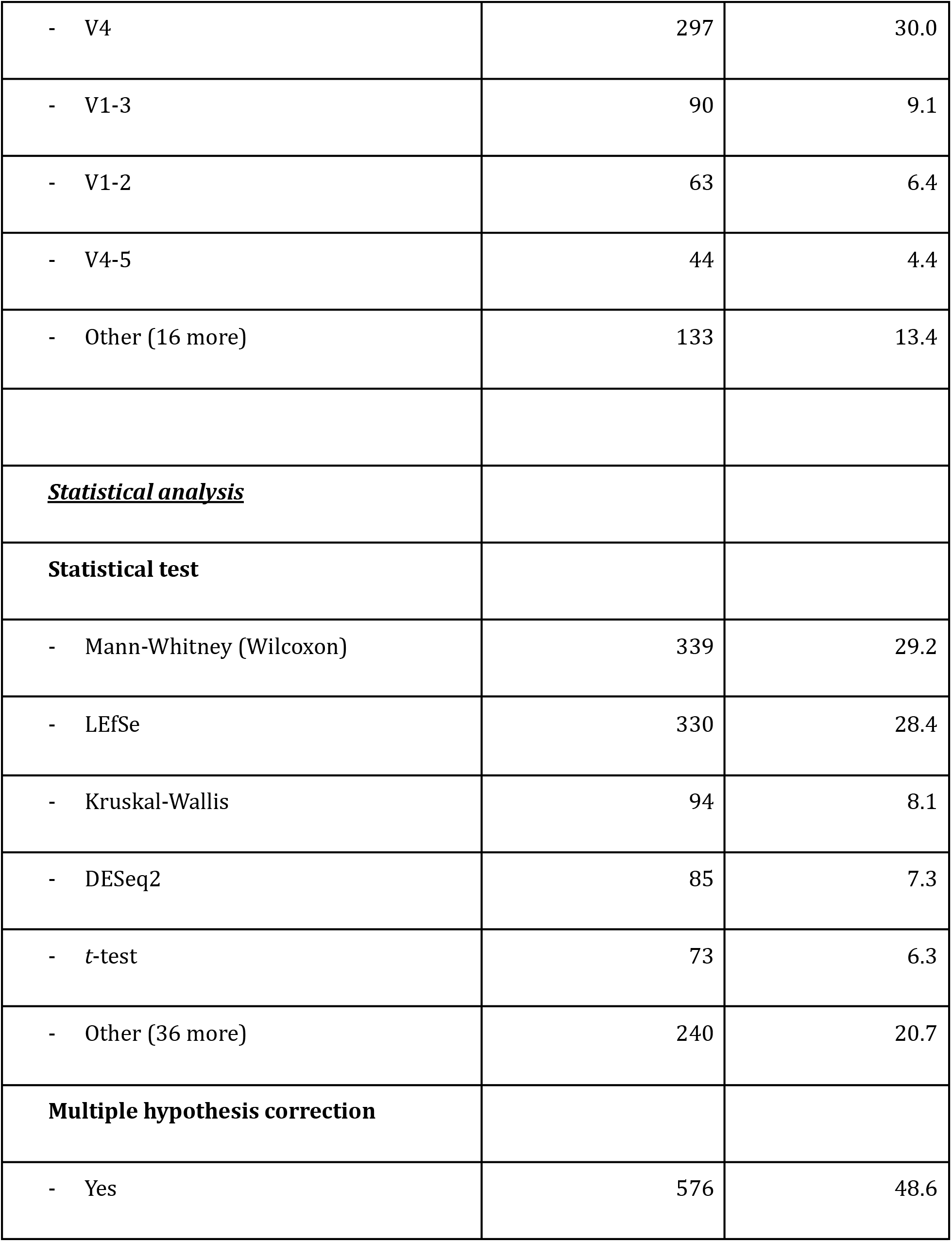
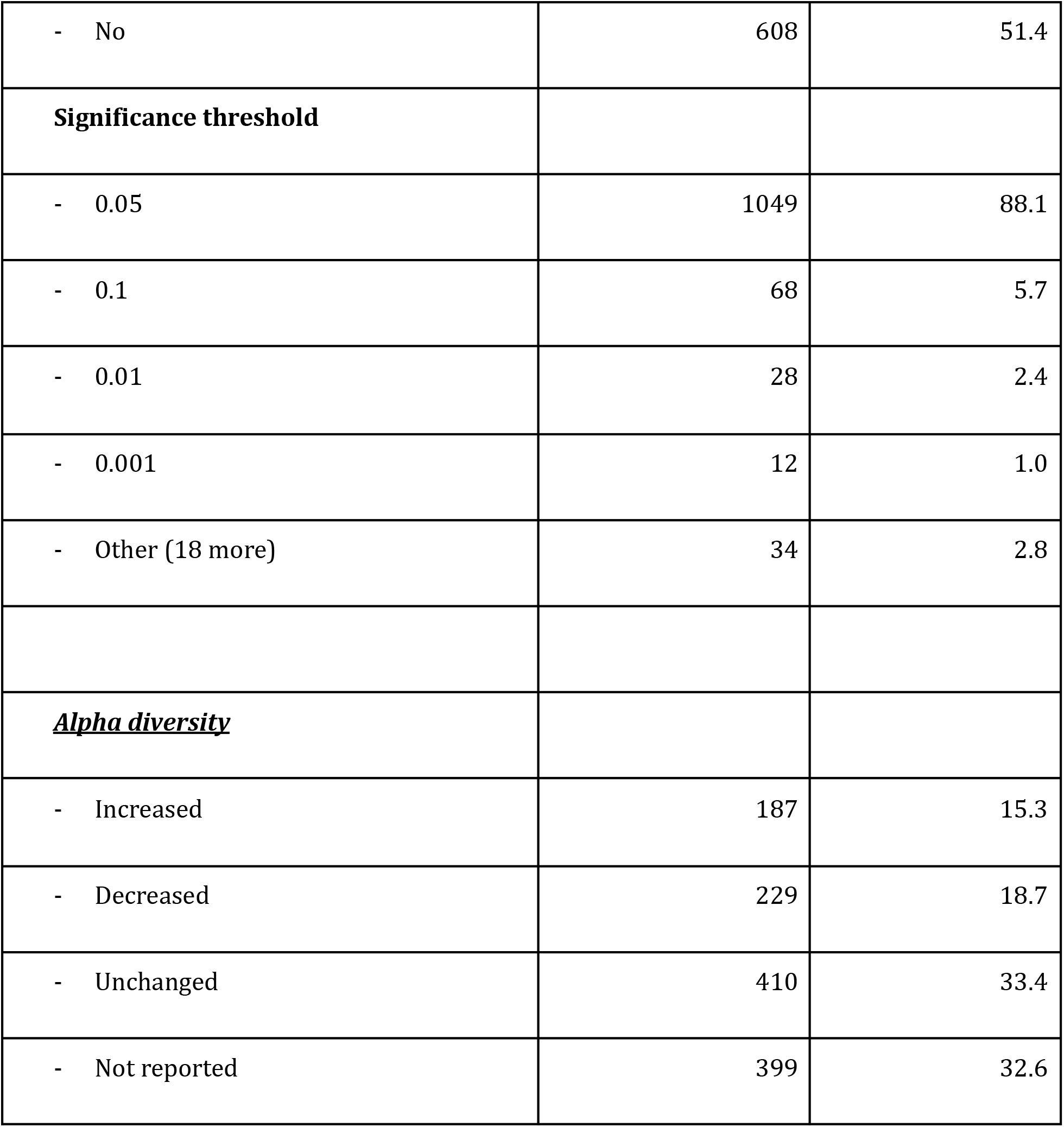
Overview of curated metadata annotations in BugSigDB. Note that the unit in the count column is the number of studies for study design, and the number of experiments for all other variables. BugSigDB defines experiments as semantic units within studies based on a defined set of characteristics about subjects, lab analysis, statistical analysis, and alpha diversity (see Methods, Section *Definition of semantic concepts*).

Most of the roughly 1,400 unique taxa contained in BugSigDB are reported as differentially abundant in fewer than 5 signatures (1,009 of 1,370 unique microbes, 69.5%, Figure 1D). Conversely, genera *Streptococcus, Prevotella, Bacteroides,* and *Lactobacillus* are each reported as differentially abundant in more than 100 signatures, reflecting the large number of species belonging to these genera and paralleling previous observations of gene signatures with certain genes being reported disproportionately often as differentially expressed^31^. Signatures contain 6 microbes on average, with roughly 55% of the signatures containing 5 or more microbes (Figure 1E).

Study conditions associated with each signature are classified according to the Experimental Factor Ontology (EFO^32^, Figure 1F), with large proportions associated with different types of cancer (23.1%), anatomical system diseases such as asthma and endometriosis (22.6%), and metabolic diseases such as obesity and diabetes (15.3%). Apart from different disease categories, substantial proportions of signatures in BugSigDB are also associated with diet (6%), use of antibiotics (4.8%), and birth delivery mode (vaginal vs. C-section, 4.1%). These distributions approximately reflect the human microbiome literature. Non-human host species remain under-represented although the database can support any host organism.

Condition-specific associations were investigated for the 10 most-reported taxa (Figure 1G). For instance, signatures containing the *Bacteroides* genus are related to metabolic disease (21%) more frequently than are all signatures (13%) (*p* = 0.003, χ^2^ = 8.4, df = 1, two-sided proportion test); *Bacteroides* is similarly enriched in signatures of antibiotic exposure (11% of *Bacteroides-*containing signatures being of antibiotics exposure vs 5% of all signatures; *p* = 0.0009, χ^2^ = 11.2, df = 1). BugSigDB makes such commonalities across groups of related studies straightforward to identify, and provides for continuous updates of these associations as the database grows.

### Curated metadata and common practices in microbiome research

BugSigDB provides curated metadata at the level of Study (study design and automatically-generated citation information), Experiments within a Study that each define one contrast for differential abundance analysis (such as characteristics about subjects, lab analysis, statistical analysis, as well as alpha diversity), Up to two Signatures within an Experiment, each of which contains one or more Taxon (see Table 1 for summary statistics of Studies and Experiments and Methods Section *Definition of semantic concepts* for details of the design). In this manuscript we use “study group” to refer to cases in case-control studies, the exposed group in exposure-control studies, and whichever group corresponds to the condition of interest with increased microbial relative abundance in other comparisons of two sample groups.

Signatures are available primarily from observational study designs: case-control (281 studies, 44.7%) and cross-sectional studies (171 studies, 27.2%) were most prevalent, while prospective cohort studies (66, 10.5%), time-series/longitudinal studies (51, 8.1%), laboratory studies (29, 4.6%), randomized controlled trials (28, 4.4%), and meta-analyses (6, 0.9%) are also present. Subject information includes host species (95.9% human out of 1,223 experiments), location, condition, body site (Figure 1), antibiotics exclusion criteria (median exclusion time = 60 days), and sample size in study and control sample groups (median total sample size = 25).

A survey of statistical methods most frequently applied for differential abundance testing in BugSigDB revealed that nonparametric tests such as Wilcoxon-family tests and the related *LEfSe* software^33^ were most frequently used, whereas recently suggested tools for differential abundance tests accounting for the compositionality of microbiome data^34^ were rarely used (Supplementary Results S1.1). Furthermore, when stratifying experiments by body site and condition, oral and vaginal samples were frequently reported with increased alpha diversity in the study group, as opposed to samples from the GI tract which were frequently found with decreased alpha diversity in the study group (Supplementary Tables S2-S4).

### Conditions with replicable microbiome changes across studies

BugSigDB facilitates meta-analysis of differential abundance studies and enables the identification of experimental conditions and disease phenotypes where microbiome changes replicate across studies. Focusing on 1,194 signatures derived from human fecal samples in 311 published studies, we computed signature similarity within conditions and assessed whether the resulting similarity exceeds the similarity of randomly sampled signatures (Figure 2). This simultaneously determines whether signatures of the same phenotype reproduce across studies, and whether different phenotypes share similar microbial signatures.

**Figure 2.**
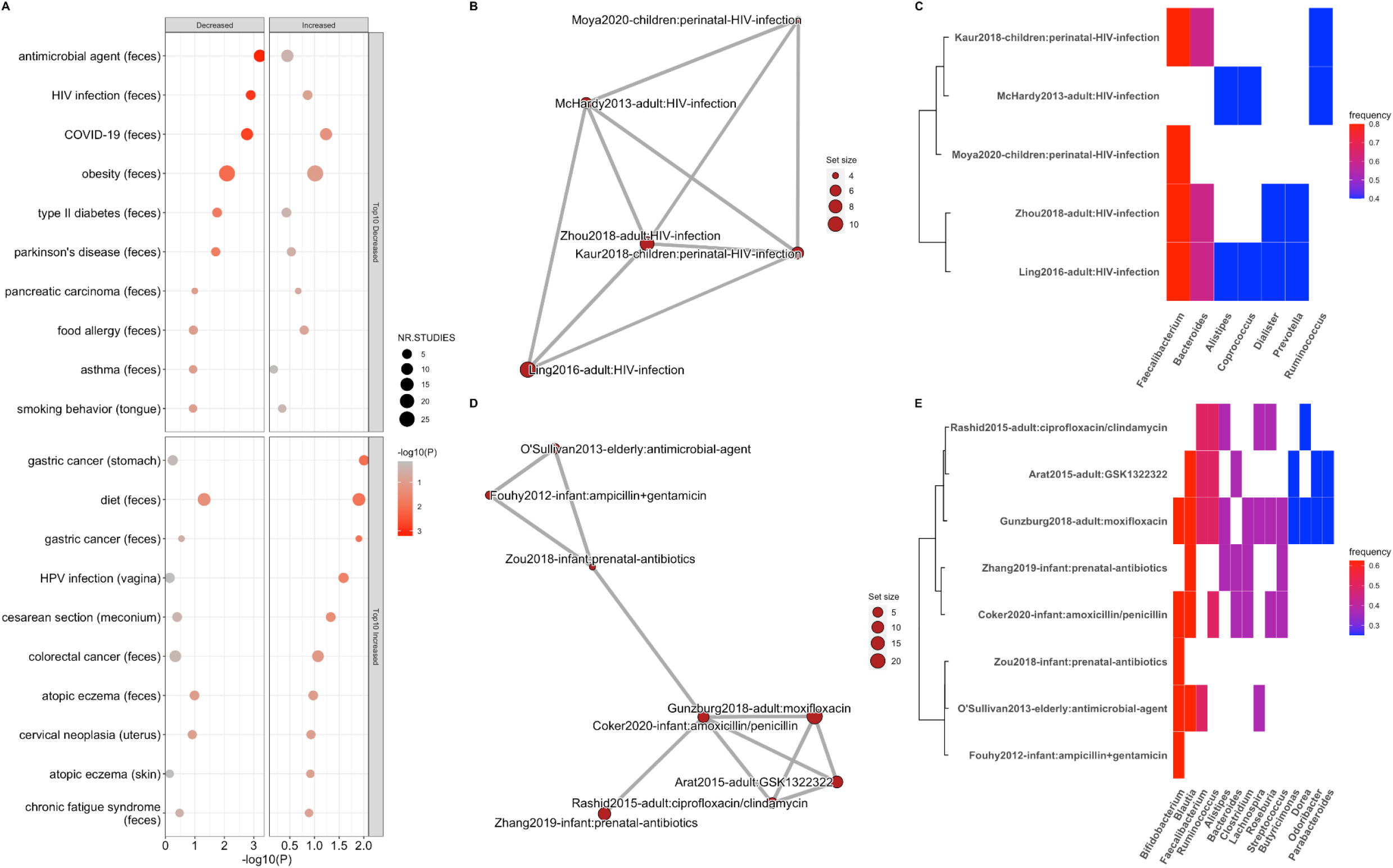
Analysis of signature similarity in BugSigDB identifies conditions with replicable microbiome changes across studies. **(A)** Signature similarity analysis for conditions from at least two studies for the same body site. Signatures were stratified by direction of abundance change into signatures with decreased abundance (Decreased panel, left) and increased abundance (Increased panel, right) in the study group. Conditions are divided into two groups: conditions with highest reproducibility for signatures of decreased abundance (Top10 Decreased panel, top) and increased abundance (Top10 Increased panel, bottom) in the study condition. Shown is the nominal *p*-value (*x*-axis, negative log scale) obtained from testing whether the semantic similarity of signatures for each condition (*y*-axis) exceeds the semantic similarity of randomly sampled signatures (one-sided resampling test). The size of each dot corresponds to the number of studies investigating a condition. **(B-E)** Example exploration of two top ranked conditions in more detail: fecal signatures of decreased abundance for patients with HIV infection (B,C), and fecal signatures of decreased abundance for patients treated with antibiotics (D,E). **(B,D)** Semantic similarity between signatures. Each node corresponds to a signature. The size of each node is proportional to the number of taxa in a signature. More similar signatures are connected by shorter and thicker edges. **(C,E)** Microbial contents of the signatures shown in panels B and D (*y*-axis) delineating the genera contained in these signatures (*x*-axis). Signatures are clustered by semantic similarity and the relative frequency of each genus across signatures is indicated by a color scale. Display of contained taxa is restricted to genera occurring in at least two signatures (full signatures are shown in Supplementary Figure S5 and S6).

To do this, we applied two alternative approaches for computing similarity between signatures: (1) the more restrictive Jaccard index^35^ based on pairwise overlaps between signatures harmonized to genus level, and (2) the more sensitive semantic similarity^36^ based on taxonomic distance between signatures of mixed taxonomic levels (see Methods, Section *Signature similarity*). Hierarchical clustering of signature similarity for both similarity measures was in good agreement, reflecting the dominance of genera reported so far in BugSigDB, but demonstrated better resolution using semantic similarity compared to the sparse results obtained from the application of Jaccard similarity (Supplementary Figure S4). The advantages of semantic similarity may grow as taxonomic ranks in BugSigDB become more mixed due to increased reporting of species-level results made possible by shotgun metagenomics.

To assess replication by independent studies of the same condition, we compared semantic similarity between signatures reported for a single condition to the similarity of randomly sampled signatures in repeated simulation, and ranked conditions based on the resulting empirical *p*-value (Figure 2A). Differential abundance signatures of antibiotic treatment and HIV infection were among the most consistent, each investigated by 5 or more studies in BugSigDB^37–48^. Closer inspection of signatures of decreased abundance after antibiotics treatment (semantic similarity = 0.64, one-sided resampling *p*-value = 0.0005, Figure 2D and E, Supplementary Figure S5) were enriched for genera of fastidious anaerobes that are often short-chain fatty acid producers, and displayed frequent loss of *Bifidobacterium* and *Blautia*, in agreement with previous reports^49^. On the other hand, signatures of decreased abundance of HIV-infected individuals versus healthy controls (semantic similarity = 0.68, resampling *p*-value = 0.002, Figure 2B and C, Supplementary Figure S6) displayed a loss of abundant members of healthy gut microbial communities as typically observed for diseases associated with GI inflammation^50^, but also resembling a response to antibiotics treatment, a likely side effect of antibiotics often being prescribed for HIV-positive patients to prevent or treat opportunistic and associated infections^51^. Additional examples of replicable microbiome shifts between studies included similarity (i) among fecal signatures of decreased abundance in patients with COVID-19, displaying alterations associated with intensive care units and antibiotics treatment^52^ (Supplementary Figure S7), and (ii) among stomach signatures of patients with gastric cancer driven by consistently increased abundance of *Streptococcus, Lactobacillus,* and *Prevotella* (Supplementary Figure S8).

### Bug set enrichment analysis of colorectal cancer signatures

We integrated BugSigDB signatures with the manually curated metagenomic datasets from curatedMetagenomicData^53^ in order to systematically benchmark enrichment methods from the `EnrichmentBrowser` package^7,54^ and evaluate whether top-performing gene set enrichment methods can be directly applied to microbiome data. We applied two enrichment methods that have performed well in previous benchmarking of gene set analysis methods^7^ - Over-Representation Analysis (ORA^11^), and Pathway Analysis with Down-weighting of Overlapping Genes (PADOG^31^) - to 10 colorectal cancer datasets from `curatedMetagenomicData`. We performed enrichment analysis of all microbiome signatures from BugSigDB simultaneously, employing as “spike-in” controls two signatures of the colorectal cancer-associated fecal microbiome, derived previously by two independent meta-analyses of individual-participant data^55,56^ from 8 of the studies included in this dataset (Figure 3). The two signatures based on meta-analysis are thus expected to be among the most enriched of all microbiome signatures: in the 8 training studies for the meta-analyses due to their shared utilization in both datasets, and in the remaining two independent studies due to the relevance and large sample size of the colorectal cancer signatures. The two meta-analysis signatures can thus also be considered robust against spurious signals from studies with small sample sizes that were not included in the analysis (Supplementary Results S3.2, Supplementary Tables S5 and S6).

**Figure 3.**
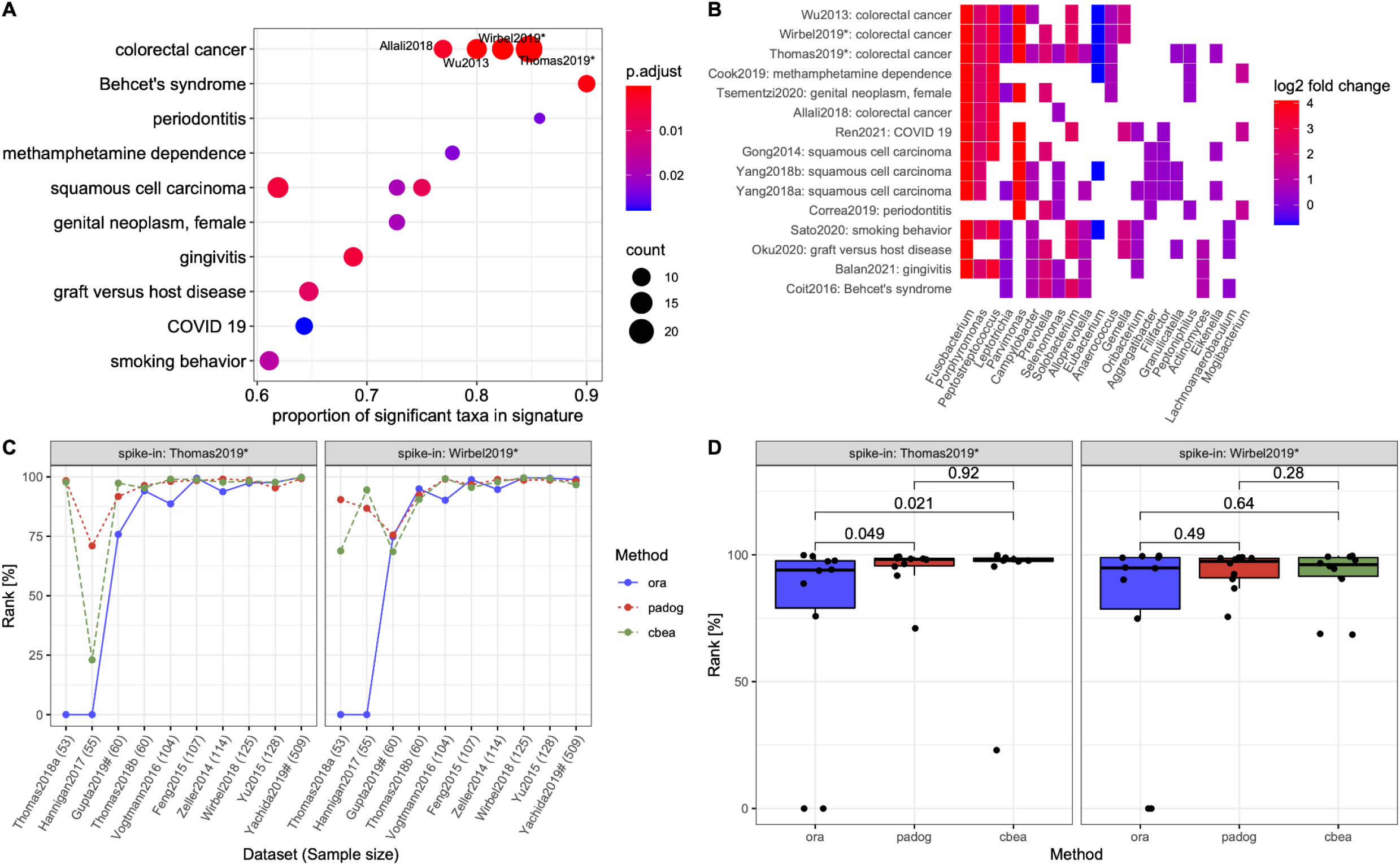
Bug set enrichment analysis reveals associations of the human microbiome with colorectal cancer and other disease phenotypes. **(A)** Overrepresentation analysis (ORA) of BugSigDB signatures with differentially abundant genera between 662 colorectal cancer samples and 653 control samples pooled from 10 published metagenomic datasets. Each dot corresponds to an enriched published signature from BugSigDB (FDR < 0.05, one-sided Fisher’s exact test, color scale). The size of each dot corresponds to the number of differentially abundant genera in a signature, given as a proportion on the *x*-axis. * Prior meta-analytic signatures from Thomas et al., 2019^55^ and Wirbel et al., 2019^56^ which reported differentially abundant species and genera from meta-analysis of 8 colorectal cancer datasets. These act as positive controls, as several of the datasets being tested for enrichment were included at the time these signatures were derived. **(B)** Differentially abundant genera (*x*-axis) in the top 15 enriched signatures (*y*-axis) from the overrepresentation analysis in (A). The *x*-axis is sorted by occurrence frequency of each genus in decreasing order, the *y*-axis is sorted by Jaccard similarity between signatures. **(C)** Percentile of ranks (*y*-axis) of both meta-analytic signatures for ORA, pathway analysis with down-weighting of overlapping genes (PADOG), and competitive balances for taxonomic enrichment analysis (CBEA) relative to all signatures when applied to the 10 published metagenomic datasets individually (*x*-axis). ^#^ Datasets not included as training sets in the meta-analyses of Thomas et al., 2019, and Wirbel et al., 2019. **(D)** Assessment of statistically significant differences of the percentile ranks on the *n*=10 independent datasets shown in (C) between the three methods using a two-sided Wilcoxon’s signed-rank test. Box plots show median (vertical line), interquartile range (IQR, box) and ±1.5× IQR (whiskers).

ORA analysis of 647 BugSigDB signatures yielded 19 signatures enriched in contrasts employing 662 colorectal cancer samples and 653 control samples from 10 datasets (FDR < 0.05, one-sided Fisher’s exact test, Figure 3A). Only signatures containing 5 or more genera associated with any condition were included. The two positive control spike-in signatures from Thomas et al., 2019^55^ and Wirbel et al., 2019^56^ were top-ranked as expected. Other enriched signatures included colorectal cancer signatures from Wu et al., 2013^57^ and Allali et al., 2018^58^. These are notable because neither study was included in the 10 datasets from which the meta-analysis signatures were computed. Additionally, both were based on 16S amplicon sequencing, whereas the meta-analysis signatures were based on shotgun sequencing. This analysis therefore also provides a proof of concept for integrating species-level signatures from shotgun metagenomic data with genus-level signatures derived from 16S amplicon profiles (see Methods, Section *Bug set enrichment analysis*), while also providing independent replication of the signatures from Thomas et al., 2019^55^ and Wirbel et al., 2019^56^.

The presence of 11 enriched signatures (58%) from saliva samples from studies of oral diseases such as gingivitis, peptic esophagitis, and oral carcinoma, is consistent with recent reports that oral to gut microbial introgression is a feature of colorectal cancer^59^, and with periodontal diseases being a well-established risk factor for CRC^60,61^. Frequently overlapping genera between the enriched signatures include *Fusobacterium*, *Porphyromonas*, and *Peptostreptococcus*, all displaying strongly increased abundance in colorectal cancer patients relative to healthy controls (Figure 3B).

Although these findings demonstrate the usefulness of ORA as a fast and effective enrichment method for microbiome signatures, the method has known shortcomings in the presence of correlated features^62^ or an inappropriately large feature universe^63^. The PADOG method, a top performer in several independent assessments^7,64,65^, is theoretically superior, as it applies sample permutation to preserve correlations and, by working on the full abundance matrix, does not require thresholding on differential abundance or the definition of a feature universe. In addition, the method downweights frequently overlapping microbes between signatures (such as those displayed in Figure 1D), leading to increased sensitivity and the identification of more specific signatures for the phenotype under investigation.

We therefore benchmarked PADOG against ORA in each of the 10 individual-participant shotgun metagenomics datasets of colorectal cancer, and compared the rankings of the two spike-in signatures from the meta-analyses of Thomas et al., 2019^55^ and Wirbel et al., 2019^56^ (Figure 3C). On average, PADOG ranked the spike-in signatures better than ORA, although the difference was statistically significant only for the spike-in signature from Thomas et al., 2019 (*p* = 0.049, two-sided Wilcoxon signed-rank test, Figure 3D). This difference was largest for datasets with smaller sample sizes, where the lack in power was more detrimental for ORA than for PADOG (Supplementary Figure S9).

Despite the apparent effectiveness of established gene set enrichment methods for application to microbiome data, these methods were not developed with microbiome data in mind^34,66^. Competitive Balances for taxonomic Enrichment Analysis (CBEA^18^) is a recent taxonomic enrichment method specifically developed for microbiome data that accounts for compositionality via application of an isometric log-ratio transformation of relative abundance data for the computation of sample-level enrichment scores. Benchmarked against ORA and PADOG in the CRC setting (Figure 3C), CBEA tended to rank the spike-in signature from Thomas et al., 2019, significantly higher than ORA (*p* = 0.021, Wilcoxon signed-rank test), but did not display a notable performance gain over PADOG (*p* = 0.92, Figure 3D).

### Microbe co-occurrence and mutual exclusivity within signatures

BugSigDB enables the exploration of compositional patterns within signatures of differential abundance for different body sites. Focusing on 1,194 signatures of fecal microbiomes from 311 published studies, we analyzed patterns of co-occurrence and mutual exclusivity for individual microbes and groups of microbes (Figure 4). Inspection of the top 20 genera most frequently reported as differentially abundant in signatures from fecal samples (Figure 4A) revealed genera predominantly belonging to the phyla *Firmicutes* (13 genera) and *Bacteroidetes* (4 genera), in agreement with those being the dominant phyla of the human gut microbiome^67,68^. Among the 20 were *Bacteroides, Prevotella,* and *Ruminococcus,* three dominant gut genera that are highly variable in relative abundance^69^ and have a large effect on gut microbiome clustering^70^.

**Figure 4.**
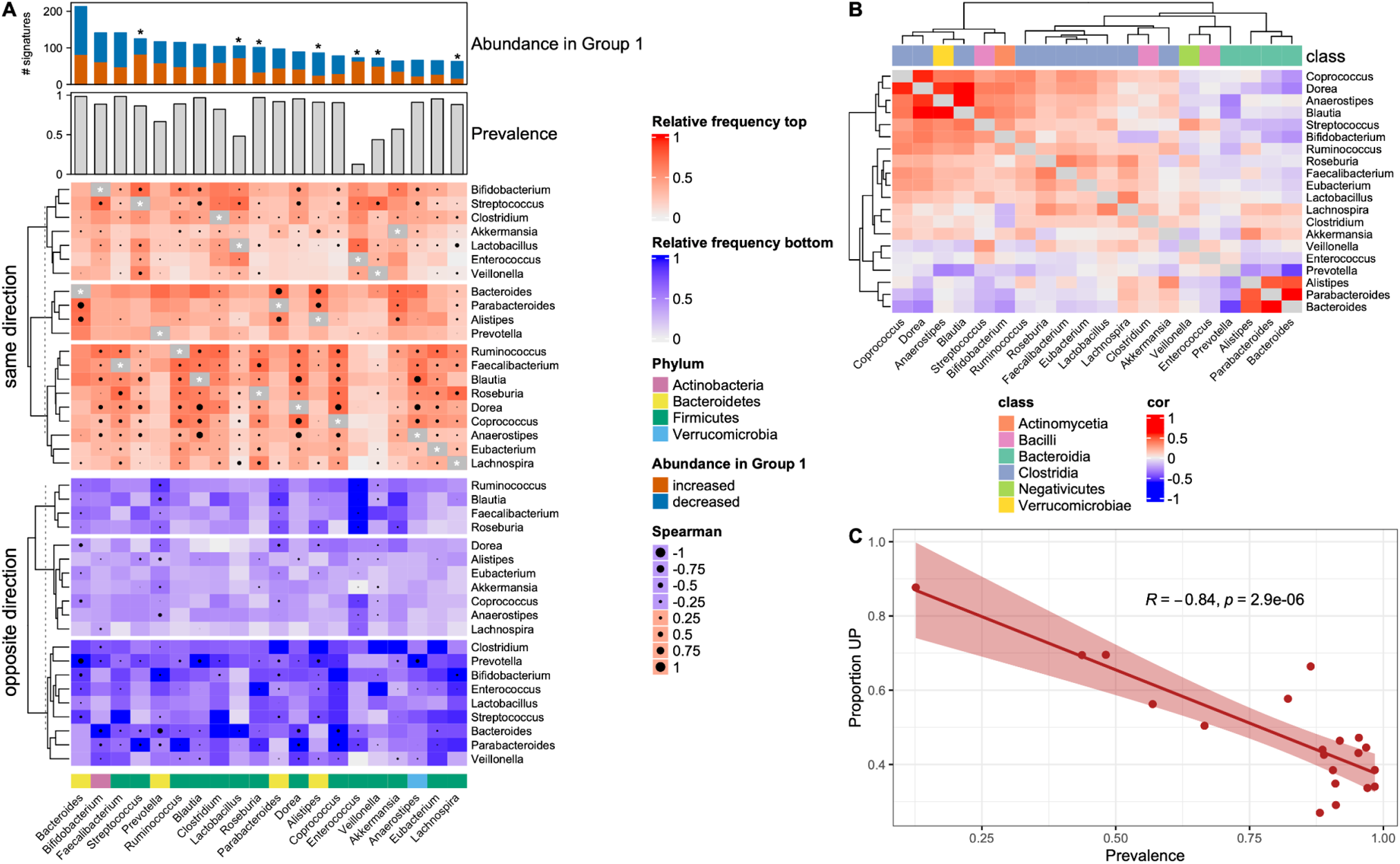
Microbe co-occurrence and mutual exclusivity in BugSigDB reveals recurrent groups of taxa within signatures of differential abundance. **(A)** Microbe-microbe co-occurrence and mutual exclusivity across 1,194 signatures of fecal microbiomes from 311 published studies. “Abundance in Group 1” shows the top 20 genera most frequently reported as differentially abundant in the study group of these signatures. Stars indicate microbes which tend to be predominantly unidirectional, i.e. reported specifically with increased or decreased abundance in the study group (based on a >2-fold difference in proportions). “Prevalence” shows the prevalence of these genera in 9,623 healthy adult stool samples pooled from 68 different studies. The top heatmap shows co-occurrence between these top 20 genera, i.e. in how many signatures these microbes are reported together as differentially abundant with the same direction of abundance change (i.e. occurring together in either the signature of increased or decreased abundance). The bottom heatmap shows mutual exclusivity between these top 20 genera, i.e. in how many signatures these microbes are reported as differential abundant with opposite direction of abundance change (i.e. one microbe in the signature of increased abundance and the other in the signature of decreased abundance, or vice versa). (**B)** Spearman correlation between the top 20 genera in healthy adult stool samples as indicated by circles of varying size in A. **(C)** Proportion of signatures where each of the top 20 genera was reported with increased abundance in the study group (*y*-axis) against the prevalence of these genera (*x*-axis) in healthy adult stool samples as shown in A. Correlation coefficient and *p*-value of a two-sided Pearson correlation test are shown at the top. The error bands indicate the 95%-confidence interval of the linear regression fit.

When comparing the proportions of signatures where these genera were reported either with increased or decreased abundance in the study sample group (panel “Abundance in Group 1” in Figure 4A), the genera most disproportionately increased in the study group were *Enterococcus* (64 out of 73 signatures, 87.7%), *Lactobacillus* (73/105, 69.5%), *Veillonella* (50/74, 69.4%), and *Streptococcus* (83/128, 66.4%). Pathogenicity of certain *Streptococcus* and *Enterococcus* species is well documented^71,72^, whereas *Lactobacillus* and *Veillonella* are typically considered to be commensal gut microbiota of limited pathogenicity^73,74^. Genera instead *decreased* in the study group were *Roseburia* (77/103, 74.8%), a beneficial gut organism with established anti-inflammatory activity^75^; and *Alistipes* (61/88, 69.3%), for which protective and harmful associations with a range of diseases have been described^76^.

To elucidate the extent to which frequency of differential abundance in disease phenotypes can be explained by prevalence in the healthy gut microbiome, we contrasted these findings with the prevalence of these genera in 9,623 stool samples from healthy adult controls of 68 different studies in `curatedMetagenomicData` (panel “Prevalence” in Figure 4A). We observed a strong negative correlation between prevalence, measured as the percentage of control samples in which the genus is observed at non-zero relative abundance, and the proportion of BugSigDB signatures in which the genus is reported with increased abundance in disease (*r* = -0.84, *p* = 3 · 10^-6^, two-sided Pearson correlation test, Figure 4C). This indicates that across the many diseases and exposures present in BugSigDB, high-prevalence genera tend to be lost in study groups relative to controls, and low-prevalence genera have increased abundance in the study group. This was particularly apparent for *Enterococcus*, a genus of low prevalence in healthy samples (13%), that was reported almost exclusively with increased abundance in the diseased group (64 out of 73 signatures, 88%). Presence of *Enterococcus*, accompanied by exclusion of other genera of the *Clostridia* class (below), may therefore be considered as a commonly reported “dysbiotic” signature.

The human gut is a complex ecosystem where microbes compete and cooperate^77^. To investigate which of these interactions are associated with disease, we next studied patterns of co-occurrence and mutual exclusivity for the top 20 genera most frequently reported as differentially abundant. For each pair of microbes we counted the number of signatures where both microbes were reported with either the same or opposite direction of change in relative abundance (top and bottom heatmap in Figure 4A). This resulted in clusters of co-occurrence driven primarily by functional and phylogenetic similarity, with frequent co-occurrence of genera of the phylum *Bacteroidetes* and blocks of positive associations in the class *Clostridia* (top red heatmap in Figure 4A, third cluster from top to bottom). On the other hand, clusters of mutual exclusivity displayed clear signs of the established *Firmicutes-Bacteroidetes* gradient in gut microbiomes^67^ and a strong negative association between *Bacteroides* and *Prevotella* within the phylum *Bacteroidetes,* as previously reported^78^ and also observed in healthy samples (*r* = -0.49, *p* < 2.2 · 10^-16^, two-sided Spearman correlation test, bottom blue heatmap in Figure 4A, third cluster from top to bottom). Overall, these patterns largely recapitulated correlation of these genera in healthy adult stool samples (Figure 4B), arguing against the existence of specific disease-promoting interactions between these genera. Exceptions were patterns of pronounced mutual exclusivity within the class *Clostridia* in signatures associated with disease, as observed for *Clostridium*, *Enterococcus*, and *Streptococcus*, which were not observed in healthy samples.

### Shared and exclusive patterns in pooled microbial signatures

BugSigDB provides opportunities for the discovery of microbial biomarkers and re-assessment of previous findings across a much larger and more heterogeneous data source than previously possible. To identify similarities between microbial shifts within and across body sites, we aggregated signatures for one body site at a time, and within body sites for one condition at a time (see Methods, Section *Signature pooling*). To account for differences in sample size between studies, we applied a voting approach where each taxon of a pooled signature obtained a weight based on the aggregated sample size of reporting studies, and performed hierarchical clustering based on pairwise similarity between the weighted meta-signatures (Figure 5). Clustering of 27 meta-signatures, representing body sites studied by at least 2 studies in BugSigDB and generated from 1,909 individual signatures of either increased or decreased differential abundance in the study group, resulted in two major body site clusters: a cluster primarily composed of oral and nasal sites, and a cluster dominated by vaginal and gastrointestinal sites (Figure 5A). This clustering was largely invariant to the similarity measure used for clustering (Supplementary Figure S10), confirming the expected dominant effect of host body site of origin, and in particular, the availability of oxygen.

**Figure 5.**
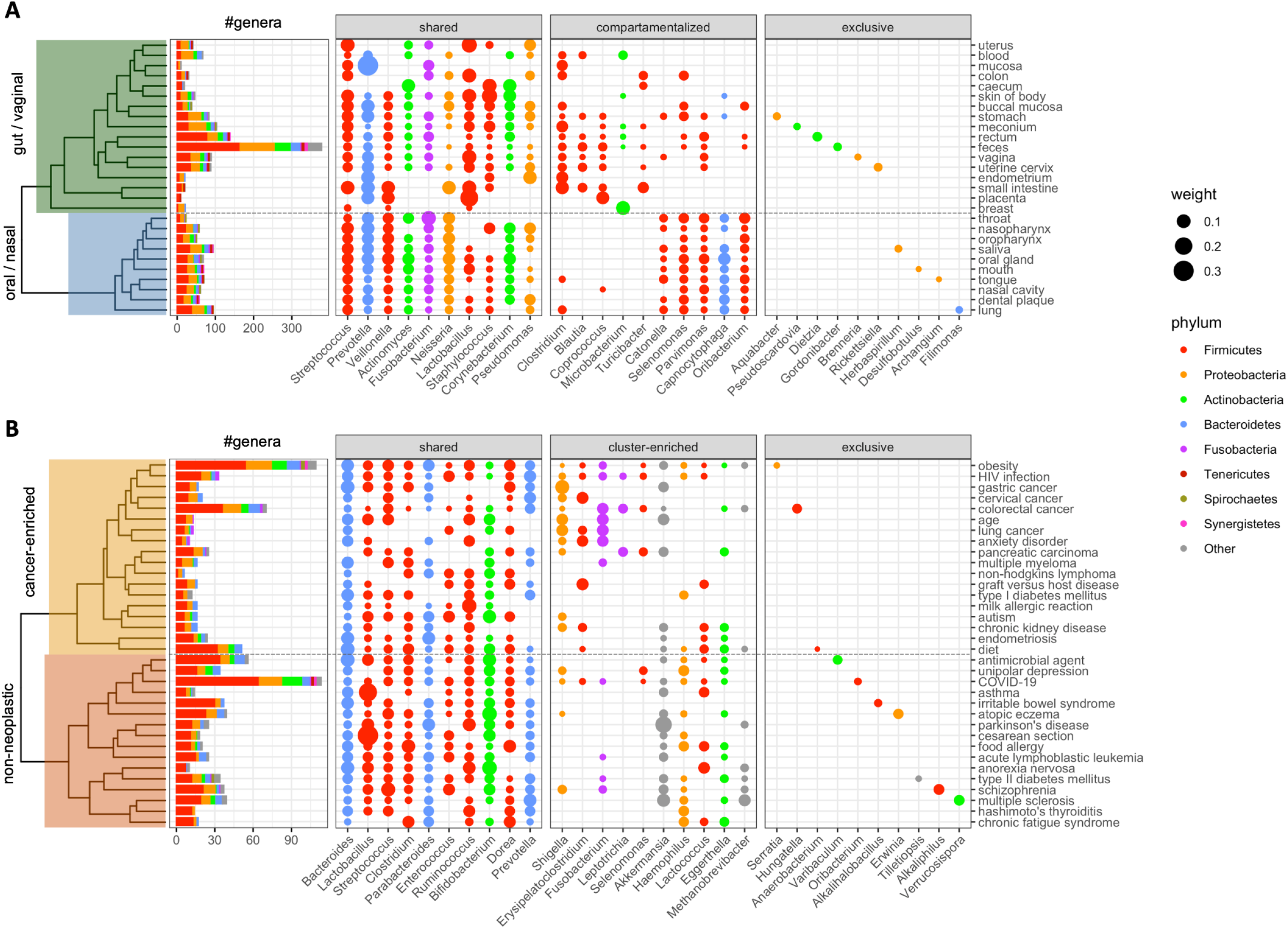
Pooled genus-level microbial signatures display robust body-site specificity and shared and exclusive patterns of gut dysbiosis between disease phenotypes. **A)** Clustering of 27 meta-signatures (*y*-axis) representing host body sites studied by at least 2 studies in BugSigDB and generated from 1,909 individual signatures. The clustering shows a separation of vaginal and gastrointestinal sites (top) from oral and nasal sites (bottom). A total of 560 different genera (*x*-axis) colored by phylum were observed across the meta-signatures. The number of genera in each meta-signature is shown in the barplot on the left. The following three panels display representative genera that frequently occur across meta-signatures (“shared”), predominantly occur in either cluster of body sites (“compartmentalized”), or were reported in only one body site as differentially abundant (“exclusive”). The size of each dot corresponds to the relative sample size of studies reporting a genus as differentially abundant. **B)** Clustering of fecal meta-signatures drawn from 34 different conditions, each represented by at least 2 studies in BugSigDB and generated from 504 individual signatures with increased abundance in the study group. The clustering shows a separation of cancer phenotypes (top) from non-neoplastic diseases (bottom). A total of 248 different genera colored by phylum were observed across the fecal meta-signatures. The number of genera in each meta-signature is shown in the barplot on the left.

Within body-site clustering of fecal meta-signatures of 34 different conditions, studied by at least 2 studies in BugSigDB and generated from 504 signatures of increased relative abundance in the study group, revealed similarities in reported differential abundance patterns between disease phenotypes (Figure 5B). This included similarities in microbial shifts for (i) HIV infection and different gastrointestinal cancers, both characterized by chronic inflammation of the GI tract and microbial signatures that point to shared pathogenic pathways including tryptophan catabolism and butyrate synthesis^79^, (ii) chronic kidney disease and autism, linked through deleterious copy number variants^80,81^ and pathogenic gut microbiota-derived metabolites produced by species of the *Clostridia* class^82,83^, and (iii) type 2 diabetes and schizophrenia, consistent with observations that people with schizophrenia are at increased risk of type 2 diabetes, and conversely, that traditional risk factors for type 2 diabetes are common in people with schizophrenia and can affect the gut microbiome, especially obesity, poor diet, and sedentary lifestyle^84^. A strong enrichment of genera of the *Clostridia* class drove the similarity between the meta-signatures of Hashimoto’s thyroiditis (12 out of 14 genera, 85.7%, *p* = 2.1 · 10^-05^, two-sided proportion test) and chronic fatigue syndrome (11/17, 64.7%, *p* = 0.003). Chronic fatigue is common in patients with Hashimoto’s thyroiditis; the disorder affects the thyroid gland, potentially also through a gut microbiota-driven thyroid-gut-axis^85^, reducing thyroid hormone production which causes extreme fatigue^86^.

Genera reported as differentially abundant exclusively in one condition, in particular those replicated by independent studies, are candidates for condition-specific biomarkers (Supplementary Figure S11, prioritized by relative sample size). This included an exclusive abundance increase of the genus *Hungatella* for colorectal cancer, which is notable given a reported role of *Hungatella hathewayi* in driving host colonic epithelial cell promoter hypermethylation of tumor suppressor genes in colorectal cancer^87^. On the other hand, an exclusive decrease in abundance was observed for *Marvinbryantia* in type 2 diabetes, for which a reduction in *Marvinbryantia* through treatment with glucagon like peptide-1 (GLP-1) receptor agonist drugs such as Liraglutide has been shown to contribute to treatment success^4^.

## Discussion

We compiled published signatures of microbial differential abundance in the BugSigDB database, assessed applicability of GSEA methods for enrichment analysis of microbial signatures, and identified common patterns of co-occurrence and mutual exclusion in differential abundance results across a broad sample of the human microbiome literature. BugSigDB is a Semantic MediaWiki that allows contribution, review, and correction by the microbiome research community and is usable through its web interface and through bulk exports compatible with all major GSEA software. It supports any taxon and any host species present in the NCBI Taxonomy, and therefore non-human hosts and studies of viromes and eukaryotes. BugSigDB has been initially seeded by ∼25 trained curators with more than 2,500 manually curated signatures from the figures, tables, main text, and supplementary material of more than 600 primary publications, providing a broadly relevant collection of machine-readable knowledge of microbial differential abundance. Manually curated metadata includes study design, geography, health outcomes, host body sites, and experimental and statistical methods. Initial analyses of the database 1) identify human diseases with the highest consistency among independently published signatures of differential abundance, 2) demonstrate the capability of established GSEA methods to prioritize CRC signatures in the analysis of individual-participant CRC datasets while adding to evidence of frequent introgression of oral pathobionts into the gut, 3) demonstrate that the prevalence of genera in fecal microbiomes of control populations is strongly correlated with being reported as decreased across diverse study conditions, 4) confirm the broad relevance of the *Firmicutes*-*Bacteroidetes* gradient in shaping common patterns of co-occurrence and mutual exclusivity in the published microbiome literature while also identifying other less dominant patterns, 5) and define sample size-adjusted consensus signatures of body sites and conditions that can simplify and clarify future analyses.

There is concern over replicability of human microbiome studies due to numerous sources of variation in complex experimental and computational quantification pipelines^88,89^. We propose an approach based on semantic similarity that can be used in systematic reviews to evaluate replication of differential abundance signatures of mixed taxonomic levels reported by independent studies, even when different laboratory methods such as 16S amplicon sequencing vs. MGX were employed. This taxonomy-aware framework provides an effective assessment of the replicability of microbiome differential abundance signatures for subsets of the literature, allowing ranking of the relative replicability of microbiome signatures consisting of different taxonomic levels across many disease phenotypes. We identified signatures associated with antibiotics treatment and chronic inflammation of the GI tract as having the highest level of consistency or replication in signatures reported by independent studies.

We pooled signatures across host body sites and experimental conditions to expand the analysis of replicability also to the ∼45% of signatures containing fewer than 5 taxa that are too small individually to be effectively compared between studies. Using a voting approach that weights each taxon by sample size of reporting studies, we constructed consensus “meta-signatures” revealing shared and specific patterns of gut dysbiosis by disease phenotype. These meta-signatures provide a framework for simplifying the interpretation of results from future studies in the context of the published literature, distinguishing specific from generic results, and informing enrichment tests by defining a universe of reported abundance changes for a body site or condition of interest.

Although single-species biomarkers are of primary interest for therapeutic interventions, they are not sufficient for capturing complex ecological patterns of co-occurrence and mutual exclusivity and interactions between microbes that may be relevant to health and disease. Inspection of published signatures in BugSigDB is an alternative approach to studying ecological patterns that complements the analysis of co-occurrence and co-exclusion in individual-participant metagenomic profiles. In agreement with previous results^53^ but across a much larger corpus of microbiome studies, we confirmed co-exclusions not specific to disease such as the phylum-level *Firmicutes-Bacteroides* gradient and the genus-level *Bacteroides-Prevotella* gradient within the *Bacteroides* phylum. On the other hand, patterns of mutual exclusivity for *Clostridium*, *Enterococcus*, and *Streptococcus* genera were specific to disease-associated signatures and not detectable in healthy samples. To distinguish between disease markers and common false positives in biomarker discovery, we compared stool signatures of disease conditions to prevalence in ∼10,000 stool specimens from healthy participants. Across the many diseases and exposures present in BugSigDB, prevalent genera in healthy fecal microbiomes tend to be reported with decreased abundance in diseased-associated fecal microbiomes. On the other hand, genera of low prevalence in healthy fecal microbiomes such as *Enterococcus,* tend to be reported with increased abundance in the disease group. Genera such as *Lactobacillus* and *Veillonella*, which are both prevalent in the stool of healthy individuals and frequently reported as increased in many study conditions, are more likely false positives or at least are not well suited as candidate biomarkers. Future work can employ stratification of BugSigDB signatures by experimental, cross-sectional, and longitudinal study designs to better infer causality.

BugSigDB enables systematic comparison of microbial signatures from new microbiome studies to previously published results. Although concepts of gene set enrichment analysis are applicable, microbiome data presents new challenges including smaller signature sizes, taxonomic relationships between features, and mixed-taxonomy signatures. We therefore benchmarked two gene set enrichment methods (ORA^11^ and PADOG^31^) that performed well in previous benchmarking^7^ and a recent taxonomic enrichment method (CBEA^18^) developed specifically for microbiome data. Valid application of gene set enrichment methods is limited to analysis at a single taxonomic level, and ORA further requires defining a realistic feature “universe” and significance threshold for differential abundance; nonetheless, all methods performed well in prioritizing signatures of CRC across numerous CRC datasets. As PADOG addresses both (i) shortcomings of ORA in the presence of inter-microbe correlation, and (ii) compositional bias in signature databases with certain taxa occurring more frequently than others, typically a result of technical or biological sampling bias^66,90^, we recommend PADOG over ORA for the routine application of enrichment analysis to microbial signatures, especially for datasets with smaller sample sizes where a lack in power typically hinders detecting individually differentially abundant features. Recently emerging microbiome-specific enrichment methods such as CBEA^18^ have the advantage of taking into account the compositional nature of microbiome data; however, they lack the independent benchmarking and implementation of major GSEA approaches. Although we found CBEA to be a sound alternative to ORA and PADOG, we did not observe notable gains of applying CBEA over PADOG in the CRC benchmark setup, indicating that basic study characteristics such as sample size and, to a lesser extent, accounting for correlation within microbial signatures, have a larger impact on identifying relevant signatures in practice than mitigating effects of compositionality.

BugSigDB is a large and diverse collection of the currently available literature on microbial differential abundance, and thus also represents certain limitations inherent to the currently available literature. Since more than 90% of the studies included in the first release of BugSigDB are based on 16S amplicon sequencing, enrichment analyses were performed at the genus level. However, some genera are functionally heterogeneous, such as streptococci, which groups deadly pathogens with common commensals and useful food-fermenting species. Species- and strain-level variations are neglected, although they can contribute to functional differences between individuals that are important in a clinical context^91,92^. With the availability of more shotgun sequencing studies in the future, it will be possible to perform enrichment analysis at higher taxonomic resolution up to the species or strain level. Furthermore, studies included in BugSigDB are heterogeneous in their design and execution, including antibiotics exclusion time frames ranging from current use to within the previous year. Restoration of baseline microbial composition following antibiotic treatment typically takes around 1 month in children and 1.5 months in adults, although several common species of the gut microbiome might take substantially longer^49,93^. Inclusion of this and other study information in BugSigDB allows further investigation and sensitivity analysis into potential sources of heterogeneity in the literature. We anticipate that broader adoption of the recently developed STORMS reporting guidelines for human microbiome studies^94^ will contribute to more efficient extraction of information from the literature for BugSigDB.

Although natural language parsing programs have potential to complement the manually curated information in BugSigDB, the majority of the curated information is too complex for currently available text mining algorithms. Natural language parsing programs typically extract patterns from unstructured text on a sentence-by-sentence basis^95,96^, but BugSigDB standardizes microbial signatures and associated experimental, epidemiological, and statistical methods from diverse Figures, Tables, Supplements, and textual descriptions that often span multiple sentences. This places many key results outside the current capabilities of text mining applications and necessitates manual curation, but improvements in machine learning, using BugSigDB as a gold-standard dataset, may enable more efficient extraction of published microbiome methods and results in the future. Moreover, automated contributions of signatures from differential abundance software via the BugSigDB API can streamline standardized reporting of results. As the community contributes additional host species and signatures of microbial physiology and morphology, BugSigDB will dynamically expand, leading to complementary insights and improvements to the systematic interpretation of microbiome studies.

## Acknowledgements

This investigation would not have been possible without the work of 58 student and intern curators who contributed data to BugSigDB: Aditi Singh, Aguobi Amara, Aiysha Shahid, Atrayee Samantha, Barakat Dindi, Busayo Ojo, Chioma Grace, Chioma Nnadi, Clare Grieve, Cindy Apiyo, Cynthia Anderson, Danya Birnbaum, Desire Oluwarotimi, Ella Jessica, Esther Ewete, Eunice Mirti, Fatima Zohra, Frans Cuevas, Ibizugbe Merit, Ifeanyi Kalu, Irem Kahveci, Jacquelyn Shevin, Jessica Shudy, Joyessa Dey, Khadijah Wuraola Amusat, Kelvin Joseph, Kweku Amoo, Lana Park, Lora Kasselman, Lucille Mellor, Madhubani Dey, Manuela Hoyos, Marianthi Thomatos, Martha Martin, Mary Bearkland, Mst Afroza Parvin, My Nguyen, Nadine Ulysse, Philippe Michael Lutete, Obafemi Blessing, Patricia Brianna, Phyu Han, Samara Khan, Shaima El Safoury, Sharmila Chunduri, Sophy, Swaibat Suleiman, Tangirul Islam, Titas Sil, Umadevi Yokeeswaran, Ufuoma Ejite, Utsav Patel, Valentina Pineda, Victoria Goulbourne, William Lam, Yaseen Javaid, Yu Wang, Zyaijah Bailey.

Research reported in this publication was supported by the National Cancer Institute of the National Institutes of Health under award numbers 5R01CA230551 (to LW, NS, CH, and HJ) and 5U24CA180996 (to LW).

## Author Contributions

L.G. and L.W. designed the study with inputs from H.J., S.D., N.S., and C.H. F.Z., R.A., S.E., and C.G. curated and reviewed data with inputs and contributions from L.G., C.M., and L.W. C.M., F.Z., R.A., C.G., and L.W. supervised student interns curating the data. I.H. implemented the semantic media wiki with inputs from L.G., C.M., F.Z., C.H., and L.W. L.G. implemented the bugsigdbr R/Bioconductor package with contributions from J.W. and inputs from C.M., S.G., and L.W. L.G. implemented the BugSigDBStats R package with contributions from P.S., S.G., and L.W., and inputs from C.M., A.R., and K.R. L.G. and J.W. implemented the BugSigDBExports repository with contributions and inputs from S.D. and L.W. J.W. released data on Zenodo with inputs from L.G. and L.W. L.G. developed and implemented methodology and performed data analysis and data visualization with inputs and contributions from R.G., E.F., S.D., N.S., C.H., and L.W. L.G. and L.W. wrote the manuscript with inputs from C.M., V.C., J.D., H.J., N.S., and C.H. All authors reviewed and approved the final version of the manuscript.

## Methods

Definition of semantic concepts (study, experiment, signature, taxon)

*Taxon:* a taxon, or taxonomic unit, is a unit of any rank (i.e. kingdom, phylum, class, order, family, genus, species, strain) designating a microbial organism or a group of microbial organisms.

*Signature:* a microbial *signature* or *set* refers here to a simple unordered list of microbial clades (*taxa*) sharing a common property or response to a study condition.

*Experiment:* BugSigDB defines *experiments* as semantic units within studies and records key characteristics about subjects, lab analysis, statistical analysis, and alpha diversity. For subjects, this includes host species, location, condition, body site, antibiotics exclusion, and sample size in study and control sample groups. To define the two sample groups that are contrasted for differential abundance, BugSigDB records the diagnostic criteria applied to define the specific condition / phenotype represented in the study group. Recorded lab analysis fields include sequencing type (16S or MGX) and sequencing platform (such as Illumina or Roche454). For 16S rRNA sequencing, the 16S variable region is also recorded. For the statistical analysis, recorded fields include (i) the statistical test or computational tool applied for differential abundance testing, (ii) whether multiple testing correction has been applied to adjust for an inflation of false positive findings, (iii) the significance threshold used to render taxa as differentially abundant, (iv) confounding factors that have been accounted for by stratification or model adjustment; and (v) factors on which subjects have been matched on in a case-control study, if applicable.

*Study:* BugSigDB collects and standardizes microbial signatures from published 16S and MGX microbiome *studies*. Studies are categorized by study design and each study is associated with a study identifier such as a PubMed ID and/or a DOI, depending on whether studies are indexed in PUBMED.

### Data entry, validation, and access

BugSigDB is implemented as a semantic MediaWiki^1^ web interface available at https://bugsigdb.org. It supports data entry, semantic validation, and web-based programmatic access to annotations for studies, experiments, signatures, and individual taxa. The semantic wiki (i) enforces data entry to follow the nomenclature of the NCBI Taxonomy Database^2^, (ii) enforces metadata annotation of signatures to follow established ontologies and controlled vocabulary for body site^3^, disease condition^4^, and type of evidence, (iii) provides an API to access all signatures, potentially filtered on taxonomy and metadata attributes, and (iv) allows commenting and error reporting on data elements and relationships. The data curation interface provides type-forward autocomplete to assist with valid data entry (including validation against the NCBI Taxonomy^2^, Experimental Factor Ontology^4^, UBERON Anatomy Ontology^3^, and administrator-defined controlled vocabulary for other fields such as statistical test and sequencing methods), to facilitate organization, filtering, and comparison of signatures. External contributions from the community, including signatures, annotations, and comments, are supported similarly to Wikipedia. Quality of contributions is controlled by tagging contributions as verified after review by a trusted editor, a method for flagging suspect entries, and the option to exclude from analysis unreviewed contributions or based on elements of study quality such as sample size, suspected contamination, paper retraction, batch effects, uncontrolled confounding, or a combination of these factors. In addition to standard semantic MediaWiki quality control tags on study level, custom methods are available for flagging taxa according to prevalence in frequently investigated host body sites, inclusion in published contamination blacklists^5,6^, and absence of known association with a host.

Signatures can be searched and browsed by study and experimental attributes, and by individual taxa, at bugsigdb.org. Bulk export of all signatures and associated metadata is available in plain-text formats for use in any programming language and software (including .csv, and the .gmt standard used by GeneSigDB^7^ and MSigDB^8^) of the current database version or as weekly and semi-annual snapshots. The companion `bugsigdbr` R/Bioconductor package (<u>bioconductor.org/packages/bugsigdbr</u>) provides advanced features such as ontology-based filtering, limitation of taxonomic level, look-up of individual signature and taxon pages, and conversion to application-centric formats. The exported files are compatible with most enrichment software and are included by default in our lab’s `EnrichmentBrowser` R/Bioconductor package^9^ to facilitate a large number of GSEA methods and visualizations. The `BugSigDBStats` R package continuously integrates with bugsigdb.org and provides weekly updated database statistics in an HTML report page (https://waldronlab.io/BugSigDBStats).

### Signature similarity

Signature similarity was computed based on two different measures: (1) Jaccard index based on pairwise overlaps between signatures harmonized to genus level, and (2) semantic similarity between signatures of mixed taxonomic levels. Pairwise calculation of Jaccard similarity for genus-level signatures was carried out using the `calcJaccardSimilarity` function of the `BugSigDBStats` package. Genus-level signatures from BugSigDB were obtained using `bugsigdbr`’s `getSignatures` function. Taxonomic clades given at a more specific taxonomic level (species or strain) were transformed by cutting the taxonomic tree at the genus level. The Jaccard index, also known as the Jaccard similarity coefficient, is defined as the size of the intersection divided by the size of the union of two input signatures *A* and *B*:

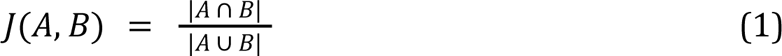

Note that by design 0 ≤ *J*(*A*, *B*) ≤ 1.

Semantic similarity was computed based on Lin’s measure of semantic similarity^10^ as implemented in the `ontologySimilarity` package from the `ontologyX` R package series^11^. Measures of semantic similarities have been proposed for comparing concepts within a taxonomy^12^, with numerous applications for biomedical ontologies^13^. Of note, semantic similarity has conceptual parallels with the computation of UniFrac distance^14^. Here, individual taxa can be considered nodes of the NCBI Taxonomy when represented as a directed acyclic graph. Computing semantic similarity between two taxa corresponds then to computing a topological similarity such as the shortest path linking the two taxon nodes. More specifically, computing Lin’s measure of semantic similarity between two taxa corresponds to computing the information content of the lowest common ancestor (LCA) of the two taxa^10^. The more frequently a taxon occurs, i.e., the higher its probability of being an ancestor of other taxa in the taxonomy, the lower its information content. If the LCA of two taxa corresponds to a taxon at a higher taxonomic level, these taxa are not very similar and this is reflected in a low information content of their LCA. Given pairwise semantic similarities between individual taxa, semantic similarity between two signatures (i.e. two taxon sets) is then obtained using a best-match average combination approach^15^, where each taxon of the first signature is paired with the most similar taxon of the second one and vice-versa.

### Bug set enrichment analysis

Metagenomic datasets providing species-level relative abundance for fecal microbiomes of colorectal cancer patients and healthy controls were obtained from `curatedMetagenomicData` 3.0^16^. Relative abundance proportions were multiplied by read depth and rounded to the nearest integer prior to obtain integer read counts compatible with bulk RNA-seq tools for differential expression and gene set enrichment analysis. For genus-level analysis, species-level counts were summed across branches using the `splitByRanks` function from the `mia` package. Given a recent assessment that reported good performance of bulk RNA-seq tools for microbiome data^17^, differential abundance analysis was carried out following the `limma`-trend approach^18^. Read counts were transformed to log counts-per-million (CPMs) using `edgeR`’s `cpm` function with a prior count of 3 to damp down the variances of logarithms of low counts^19^. Genus-level and species-level microbial signatures from BugSigDB were obtained using `bugsigdbr`’s `getSignatures` function. To keep signatures meaningfully sized, taxonomic clades given at the species-or strain level were transformed by cutting the taxonomic tree at the genus level. ORA and PADOG were carried out as implemented in the `EnrichmentBrowser` package^9^. CBEA was carried out as implemented in the `CBEA` package^20^.

### Taxon co-occurrence

Genus-level signatures from BugSigDB were obtained using the `getSignatures` function of the `bugsigdbr` package. Signatures were filtered by body site for fecal samples and stratified by direction of abundance change (increased / decreased). The top 20 most frequently occurring genera in the resulting signatures were reported. Prevalence of these genera was computed as percentage of healthy adult stool samples in which the genus was observed at non-zero relative abundance in metagenomic datasets from `curatedMetagenomicData` 3.0^16^. The correlation between prevalence in healthy samples and the proportion of signatures with increased abundance for the top 20 genera was assessed using Pearson’s correlation test as implemented in the `cor.test` function of the `stats` package. Taxon co-occurrence in signatures associated with disease was contrasted against Spearman rank correlation of the top 20 genera in healthy samples using the `cor` function of the `stats` package.

### Signature pooling

Signatures were pooled for one body site at a time, and within body sites for one condition at a time, as implemented in the `getMetaSignatures` function of the `bugsigdbr` package. Taxa within a pooled signature were weighted based on the aggregated sample size of the studies that reported this taxon as differentially abundant, divided by the total sample size of studies contributing to the pooled signature. Resulting weighted meta-signatures were clustered by semantic similarity, where the weights were incorporated into the best-match average combination approach^13^ as implemented in the `weightedBMA` function of the `BugSigDBStats` package. Analysis was restricted to body sites and conditions studied by at least two studies in BugSigDB and containing at least 5 taxa in the resulting pooled signature. Robustness of the clustering was evaluated by comparing to rank-biased overlap^21^.

### Data Availability

BugSigDB is available via a Semantic MediaWiki web interface at https://bugsigdb.org, under open-source and open-data licenses described at https://bugsigdb.org/Project:About. Weekly and semi-annual snapshots are provided in plain text file formats at https://github.com/waldronlab/BugSigDBExports for cross-language and cross-application compatibility; unprocessed snapshots are available as csv files at https://bugsigdb.org/Help:Export. The companion bugsigdbr R/Bioconductor package provides advanced data manipulation, including ontology-aware and taxonomy-aware features (https://bioconductor.org/packages/bugsigdbr). The NCBI Taxonomy database is available at https://www.ncbi.nlm.nih.gov/taxonomy. The Experimental Factor Ontology is available at https://www.ebi.ac.uk/efo. The UBERON Anatomy Ontology is available at https://www.ebi.ac.uk/ols/ontologies/uberon.

### Code Availability

Source code and open issue tracking are provided at https://github.com/waldronlab/BugSigDB. Statistical analysis was carried out using R^22^and Bioconductor^23^ and is reproducible using the code provided on GitHub^24^.

