## Supplemental materials for "BugSigDB captures patterns of differential abundance across a broad range of host-associated microbial signatures"

### S1 Supplementary Results

#### S1.1 Curated metadata in BugSigDB reveals common practices in human microbiome research

BugSigDB provides curated metadata that enable stratification of microbiome signatures by study design, sample size, and evidence type (Table 1 of the main manuscript). Recorded lab analysis fields include sequencing type (16S: 92.5%; metagenomic shotgun (MGX): 7.5%) and sequencing platform (Illumina: 68%; Roche 454: 15%; Ion Torrent: 7.2%; RT-qPCR: 6.6%). Most of the 16S studies amplified the V4 region, which has implications on which taxonomic clades can be reliably detected [1, 2].
A side benefit of the database is a survey of the popularity of different statistical tests in the published literature. Non-parametric tests for testing for differences in mean microbial abundance between two sample groups (Mann-Whitney U test, 29.2%) or more than two groups (Kruskal-Wallis rank test, 8.1%) were most frequently used, often performed using the popular `LEfSe` tool (28.4%) for metagenomic biomarker discovery [3]. Considerable fractions also employed parametric tests based on the raw read counts (via `DESeq2` [4], 7.3%) or relative abundance (using a $t$-test, 6.3%) for differential abundance testing. Recently suggested tools for differential abundance tests accounting for the compositionality of microbiome data [5] were rarely used.
As differential abundance of individual microbes can also be a side effect of systematic differences in alpha diversity between the contrasted sample groups, BugSigDB records whether and which measures of alpha diversity were reported. For most experiments, alpha diversity was either unchanged (410, 33.4%) or not reported (399, 32.6%), and roughly equal numbers of experiments reported either increased (187, 15.3%) or decreased (229, 18.7%) alpha diversity in the study group. Most frequently reported measures of alpha diversity were Shannon diversity (reflecting number of species and their relative abundance, 38.3%) and richness (number of species, 22.4%, Supplementary Table S2).

#### S1.2 Enrichment analysis of individual CRC studies from curatedMetagenomicData in BugSigDB

The individual CRC studies from `curatedMetagenomicData` (cMD) were not included in BugSigDB at the time of writing the manuscript and creating Figure 3 of the main manuscript. For reproducibility, all analyses presented in the manuscript have been carried out based on the BugSigDB v1.0.2 release (Jan 25, 2022). Figure 3A of the main manuscript reports the results of an over-representation analysis of BugSigDB signatures in the set of differentially abundant genera obtained from comparing fecal metagenomes of pooled cohorts of 662 CRC patients and 653 healthy controls from 10 cMD datasets. The two meta-analytic signatures, which are themselves derived from large pooled cohorts and are expected to report robust CRC vs. healthy abundance changes, have thus been included as spike-in / positive control signatures. The individual cMD studies that have small sample sizes are anticipated to also report spurious signatures. Given the relationship between sample size and ranking of the spike-in signatures shown in Figure 3C of the main manuscript, one would not necessarily expect all signatures derived from the individual datasets to be strongly enriched, and some might simply not report a sufficient number of differentially abundant taxa to be included in the enrichment analysis.
During the review phase of this manuscript, we confirmed this by repeating the analysis with a more recent snapshot of BugSigDB (`b87f34e`, Jan 29, 2023) which added signatures from 4 of the individual CRC studies in `curatedMetagenomicData` (Supplementary Table S5). Two of these studies (ZellerG_2014 and VogtmannE_2016) reported signatures that were too small to be included in the over-representation analysis which required a minimum of 5 genera in a signature. The other two studies comprised a medium-sized dataset (FengQ_2015, 41 CRC vs 55 healthy samples) and a large dataset (YachidaS_2019, 258 CRC vs 251 healthy samples). The resulting ranking of the signatures of FengQ_2015 and YachidaS_2019 in an

over-representation analysis of 776 BugSigDB signatures is shown in Supplementary Table S6.
In agreement with the anticipated effect of sample size, the signature from YachidaS_2019 was strongly enriched and near the top of the ranking (10 differentially abundant genera out of 13 genera total in the signature, Benjamini-Hochberg adjusted $p$-value $2.5 \cdot 10^{-5}$), whereas the signature from FengQ_2015 did not show a strong enrichment (4 differentially abundant genera out of 10 genera total in the signature, Benjamini-Hochberg adjusted $p$-value 0.16).

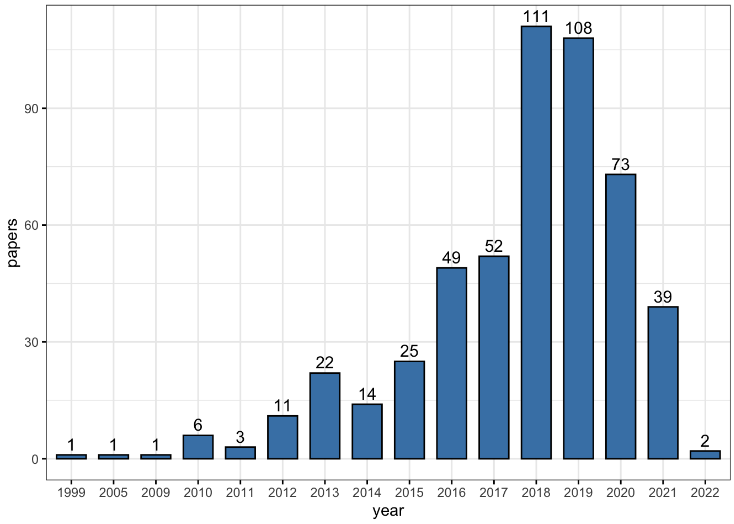

**Figure S1: Publication date of curated papers.** The curated papers cover two decades of human microbiome research, with the majority of studies being published in the last 5 years (385 / 526 studies, 73.2%).

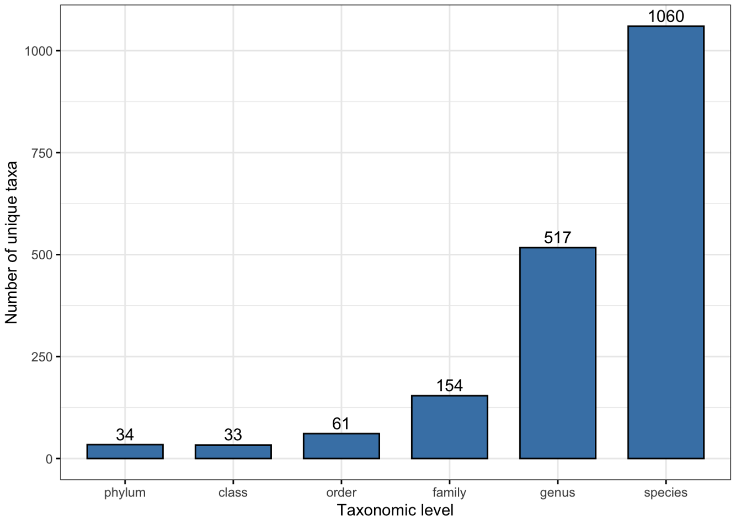

**Figure S2: Distribution of taxonomic levels in BugSigDB signatures.** Shown is the number of unique taxa ($y$-axis) for each taxonomic level on the $x$-axis.

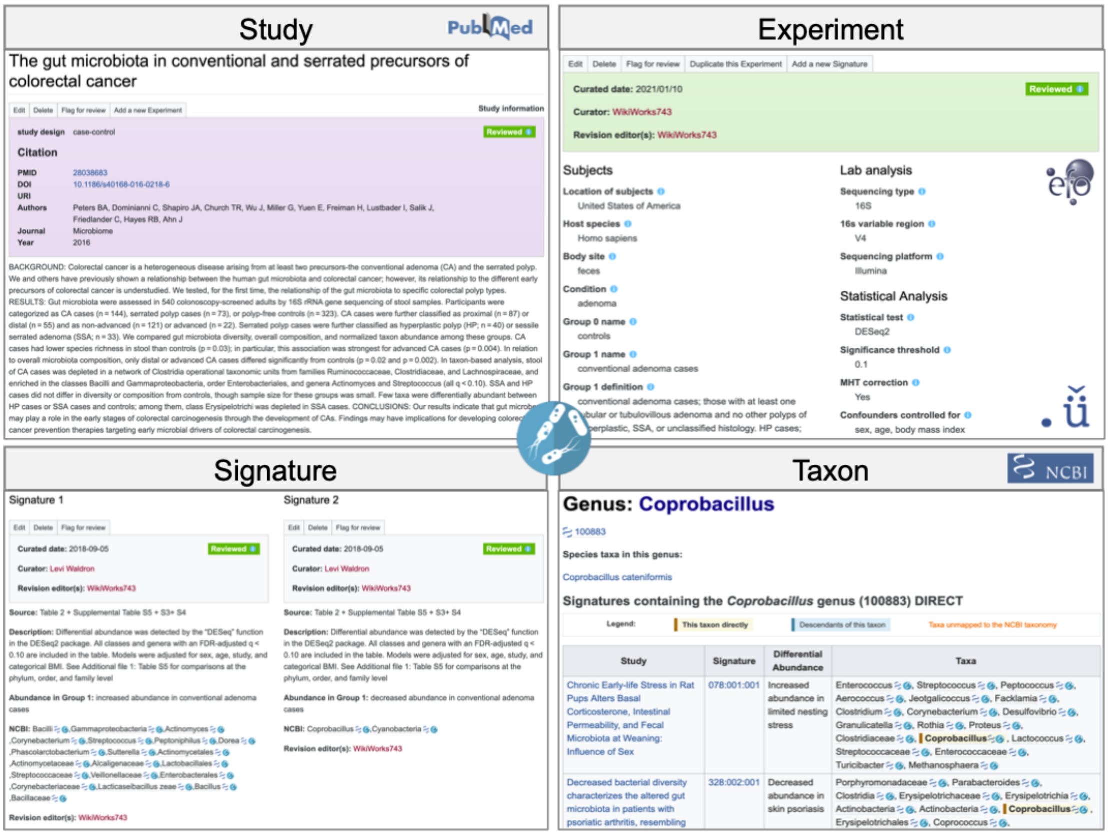

**Figure S3: A semantic MediaWiki for microbial annotations.** Semantic MediaWiki web interface available at https://bugsigdb.org for data entry, semantic validation, and web-based programmatic access to annotations for individual microbes and microbe signatures. This includes dedicated pages for (a) studies annotated with study design and linked to PubMed, (b) experiments describing characteristics of enrolled study participants, experimental procedures, and statistical analysis based on established ontologies and controlled vocabulary for body site and disease condition, (c) signatures specifying curation source and direction of abundance change (increased / decreased) of (d) individual taxa annotated following the nomenclature of the NCBI Taxonomy Database, facilitating also direct comparison of entered signatures to existing signatures in the database.

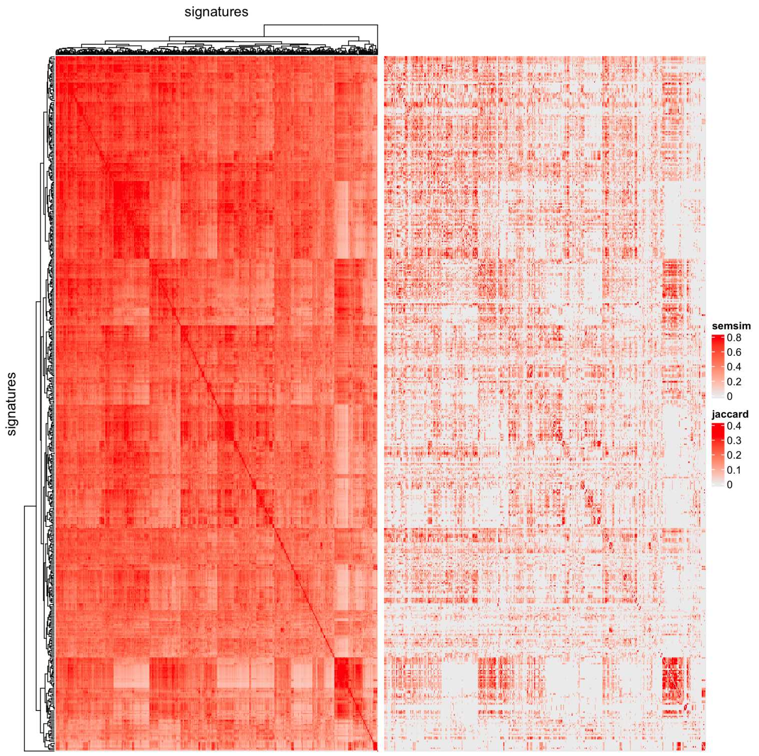

**Figure S4: Comparison of semantic similarity and Jaccard similarity.** We applied two different approaches for computing similarity between signatures: (1) the more restrictive Jaccard index based on pairwise overlaps between signatures harmonized to genus level (right panel), and (2) the more sensitive semantic similarity (left panel) based on taxonomic distance between signatures of mixed taxonomic levels (see Methods, main manuscript). Hierarchichal clustering of signature similarity for both similarity measures was in good agreement, but demonstrated better resolution of semantic similarity compared to the sparse results obtained from the application of Jaccard similarity.

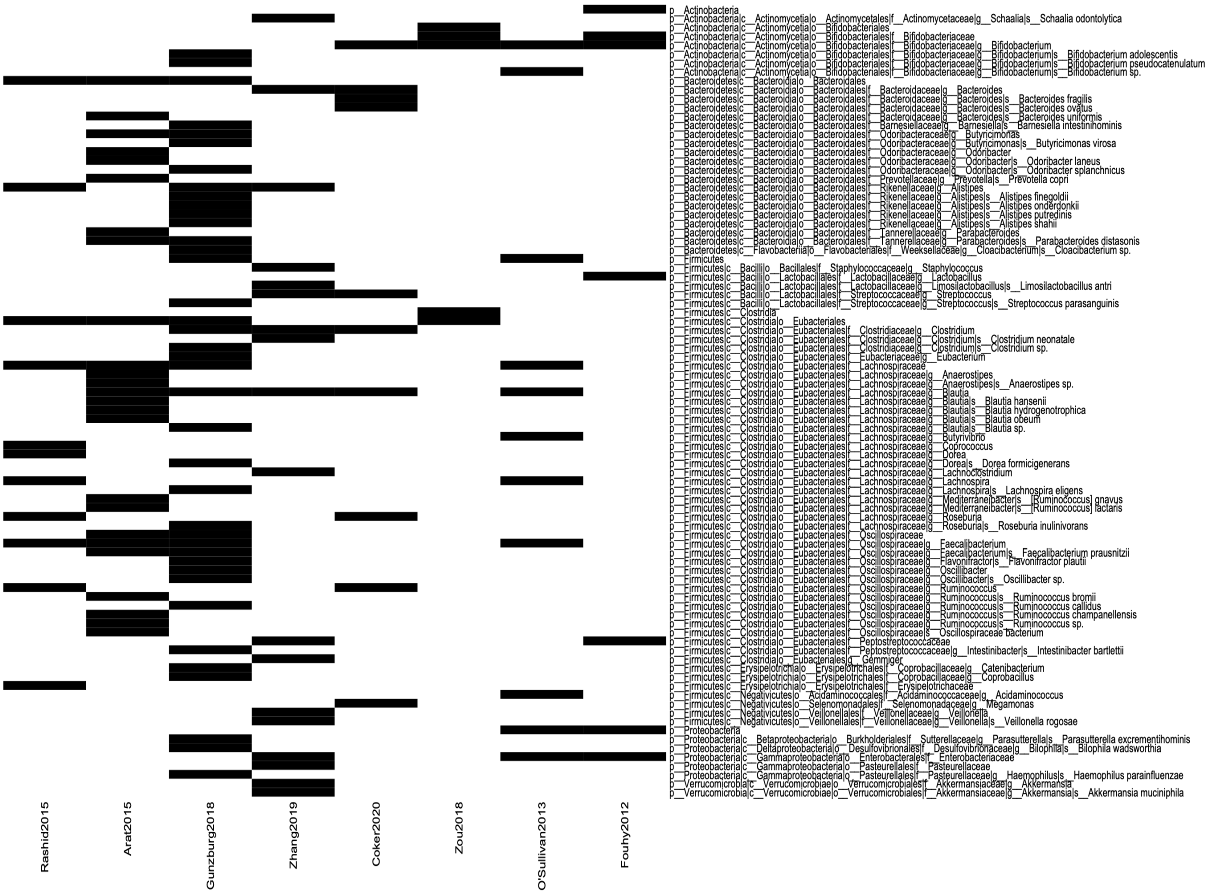

**Figure S5: Similarity between fecal signatures of decreased abundance after antibiotics treatment.** Microbial contents of the signatures shown in Figure 2D and E of the main manuscript ($x$-axis) delineating the taxa contained in these signatures ($y$-axis). Signatures contain mixed taxonomic levels from phylum to species, which is taken into account through the computation of semantic similarity between signatures.

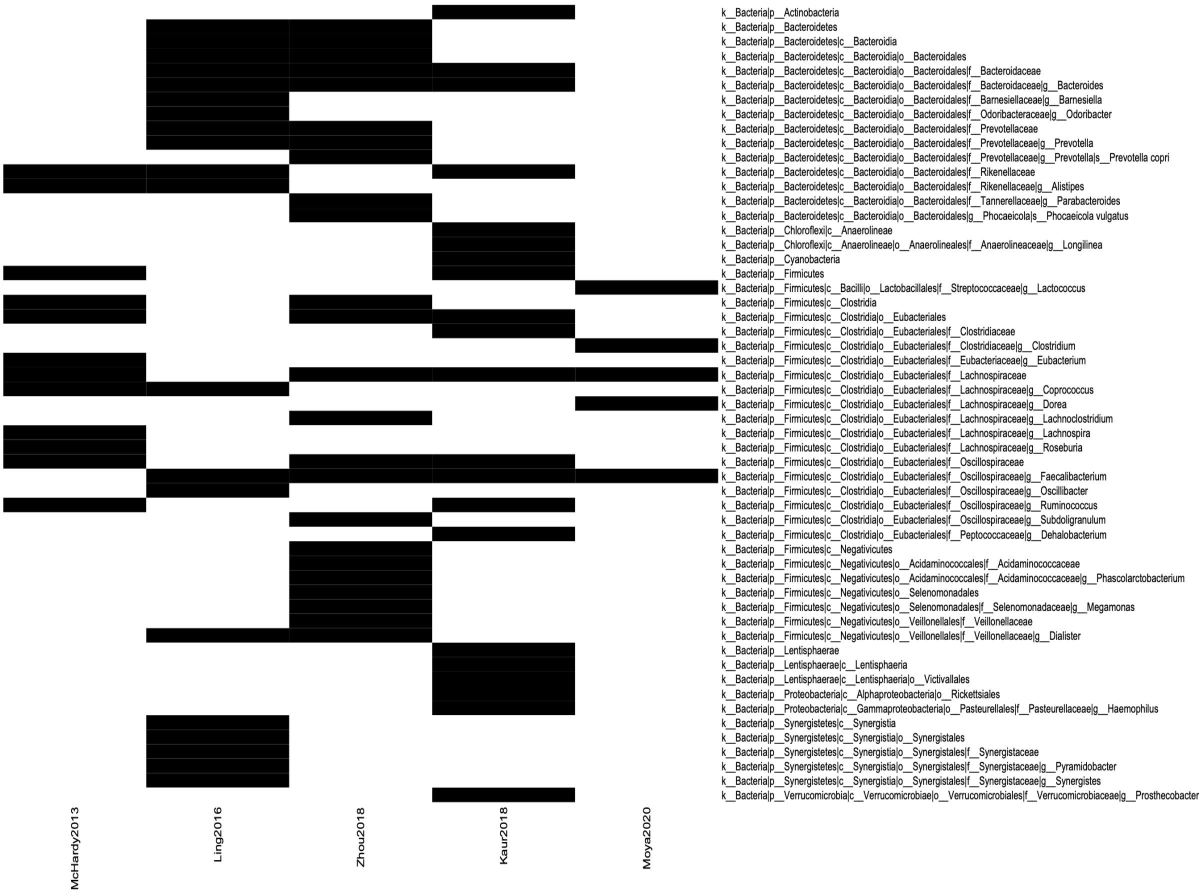

**Figure S6: Similarity between fecal signatures of decreased abundance in HIV infection.** Microbial contents of the signatures shown in Figure 2B and C of the main manuscript ($x$-axis) delineating the taxa contained in these signatures ($y$-axis). Signatures contain mixed taxonomic levels from phylum to species, which is taken into account through the computation of semantic similarity between signatures.

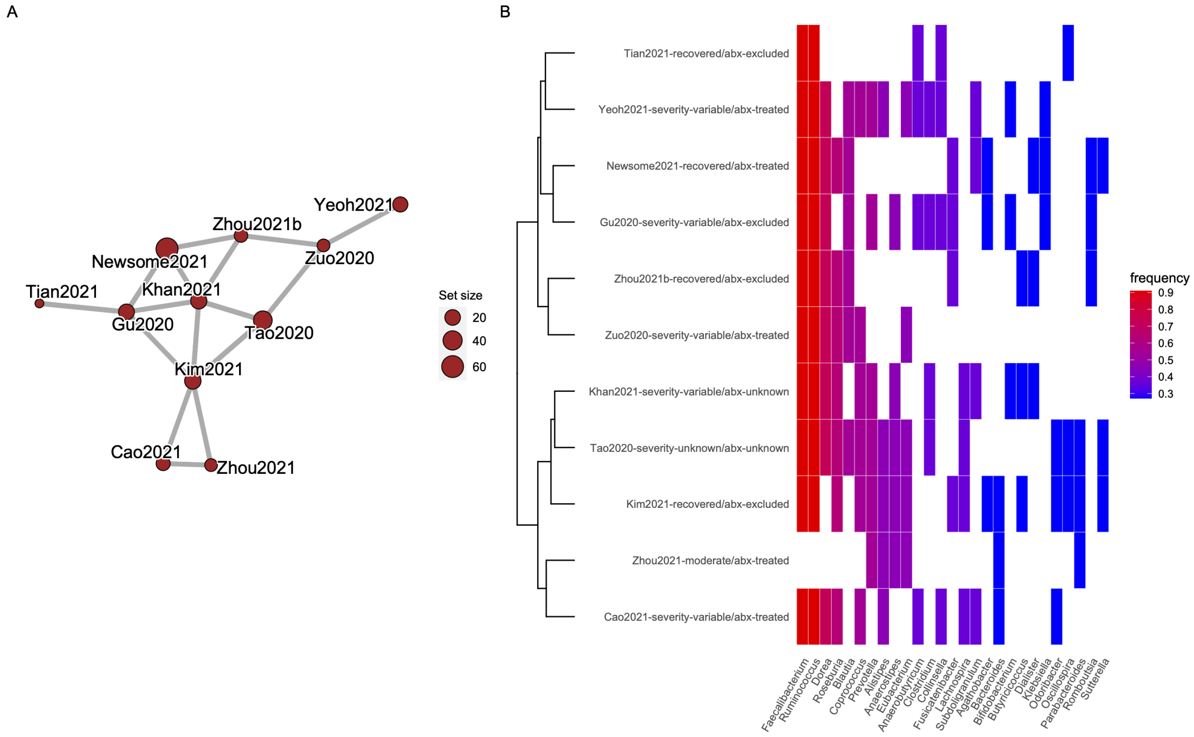

**Figure S7: Similarity between fecal signatures of decreased abundance in COVID-19. (A)** Semantic similarity between signatures. Each node corresponds to a signature. The size of each node is proportional to the number of taxa in a signature. More similar signatures are connected by shorter and thicker edges. **(B)** Microbial contents of the signatures ($x$-axis) delineating the taxa contained in these signatures ($y$-axis). COVID-19 severity and antibiotics (abx) treatment is indicated in the signature name.

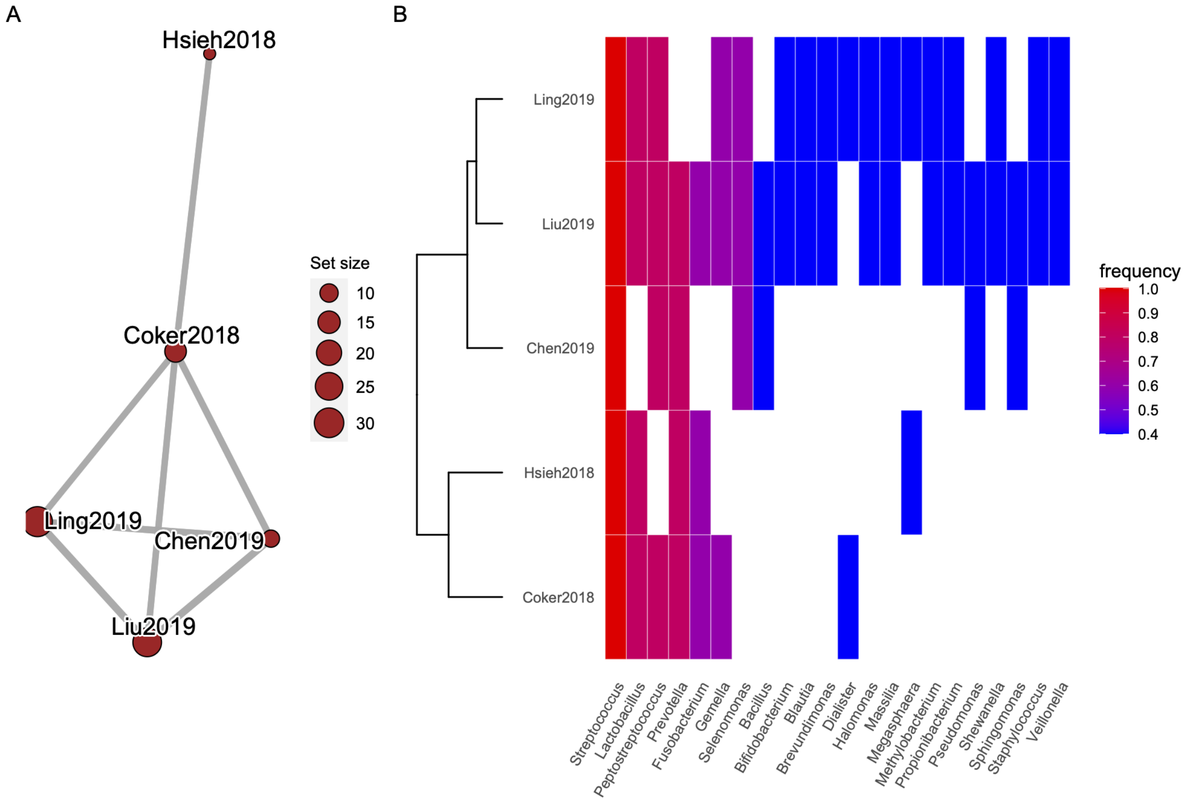

**Figure S8: Similarity between stomach signatures of increased abundance in patients with gastric cancer.** **(A)** Semantic similarity between signatures. Each node corresponds to a signature. The size of each node is proportional to the number of taxa in a signature. More similar signatures are connected by shorter and thicker edges. **(B)** Microbial contents of the signatures ($x$-axis) delineating the taxa contained in these signatures ($y$-axis).

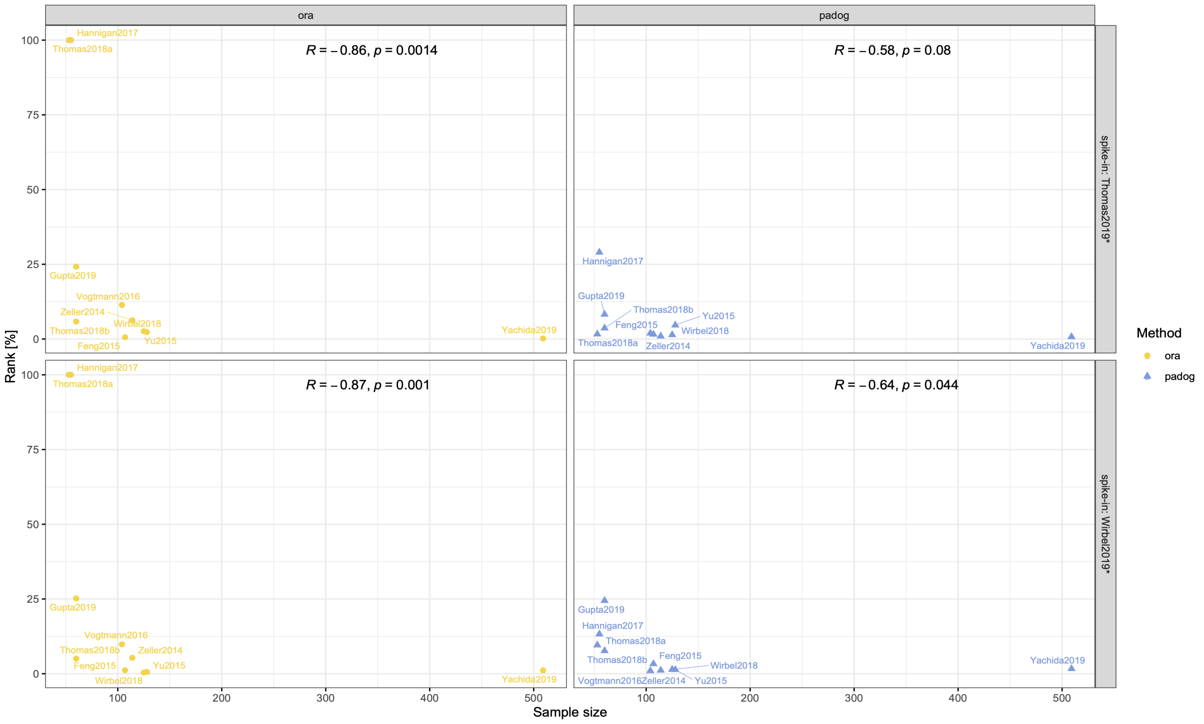

**Figure S9: Relationship between sample size and ranking of spike-in signatures.** Relative ranks ($y$-axis) of both spike-in signatures for ORA and PADOG when applied to 10 published metagenomic datasets of varying sample size ($x$-axis). The correlation and $p$-value of a two-sided Spearman's correlation test is annotated to each panel. A general trend of better ranking of the spike-in signatures for larger sample sizes is apparent for both methods, although the impact of lack in power for smaller sample sizes is stronger for ORA.

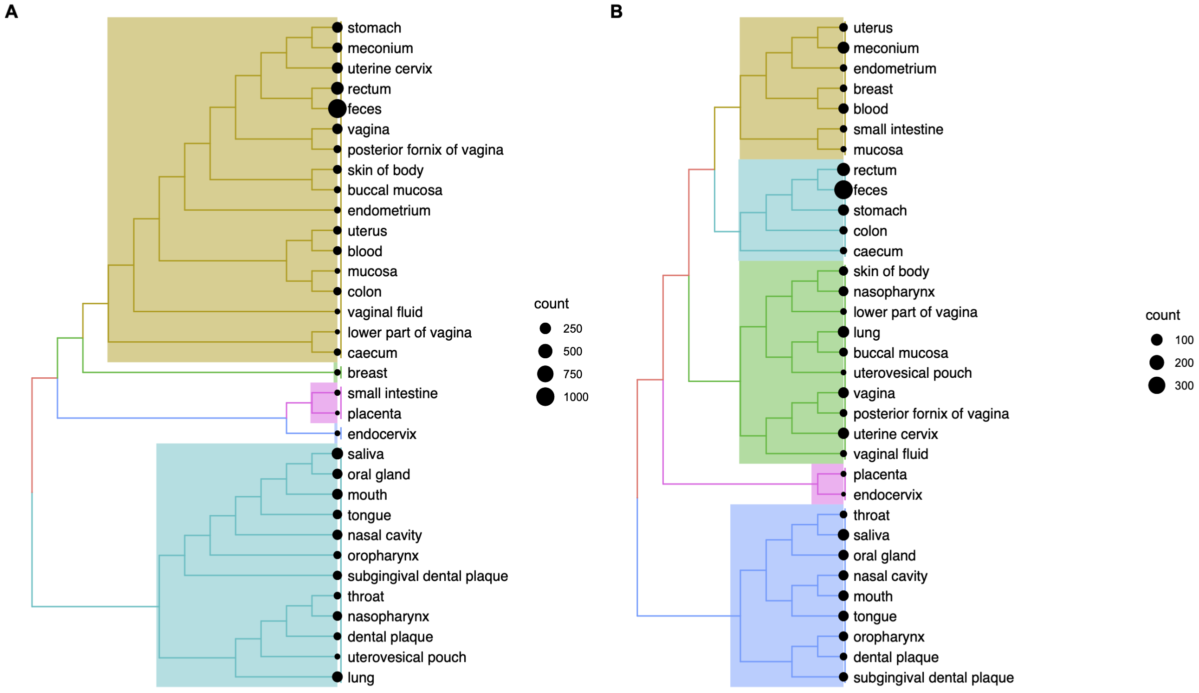

**Figure S10: Clustering of consensus signatures for body site. (A)** Clustering of weighted meta-signatures containing mixed taxonomic levels by semantic similarity [6], and **(B)** Clustering of weighted genus-level signatures by rank-biased overlap [7].

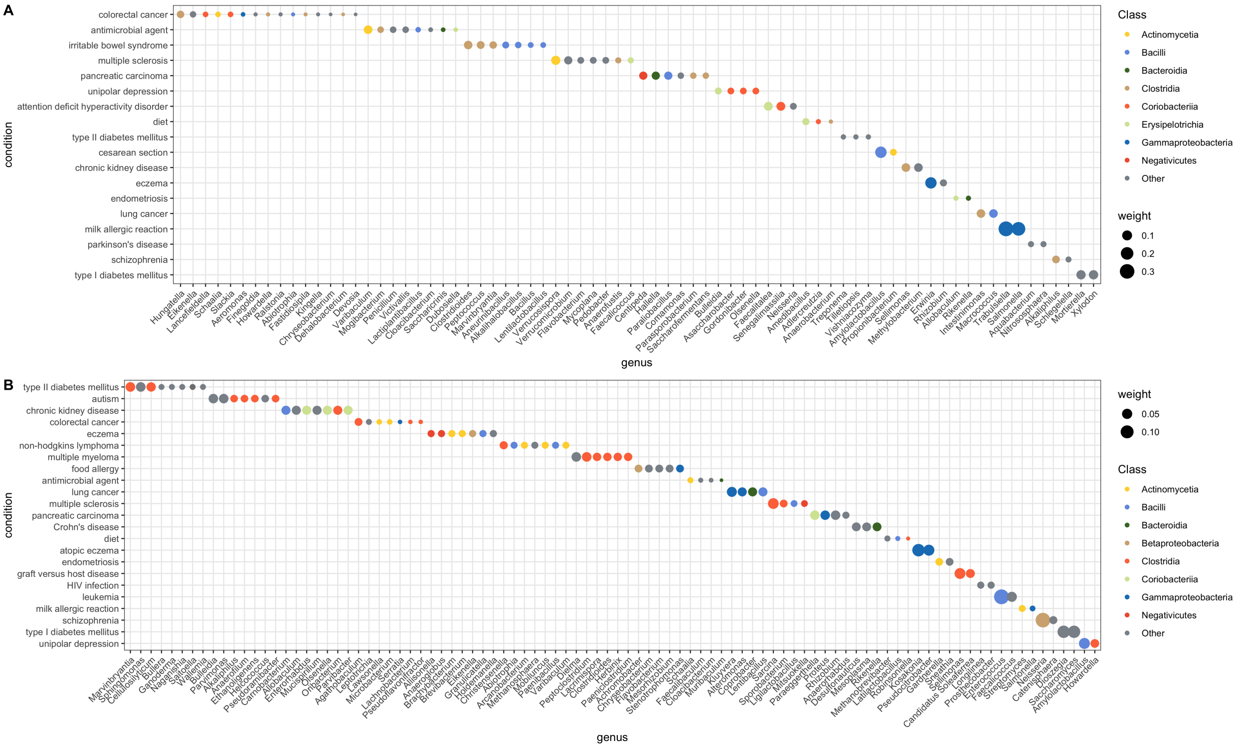

**Figure S11: Genera with mutual exclusive abundance changes between conditions.** Shown are the genera ($x$-axis) that are exclusive to one of the conditions on the $y$-axis, i.e. genera that are only reported for this condition with **(A)** increased or **(B)** decreased abundance in the study group. The weight / size of the dot corresponds to the relative sample size reporting this genus in a meta-signature for the corresponding condition.

**Table S1: Body areas and anatomical sites.** Microbiome studies in BugSigDB investigate microbiome samples from 14 broad body areas comprising more than 60 refined anatomical sites standardized based on the UBERON Anatomy Ontology [8].

| Body area | Anatomical site | UBERON ID |
|---|---|---|
| Oral | Mouth | UBERON:0000165 |
| | Lower lip | UBERON:0001835 |
| | Buccal mucosa | UBERON:0006956 |
| | Oral cavity | UBERON:0000167 |
| | Oral opening | UBERON:0000166 |
| | Oropharynx | UBERON:0001729 |
| | Pharyngeal mucosa | UBERON:0000355 |
| | Saliva | UBERON:0001836 |
| | Tongue | UBERON:0001723 |
| | Gingiva | UBERON:0001828 |
| | Internal cheek pouch | UBERON:0013640 |
| | Dental plaque | UBERON:0016482 |
| | Subgingival dental plaque | UBERON:0016484 |
| | Throat | UBERON:0000341 |
| | Hypopharynx | UBERON:0001051 |
| Nasal | Nose | UBERON:0000004 |
| | Nasal cavity | UBERON:0001707 |
| | Nasopharynx | UBERON:0001728 |
| Respiratory tract | Lung | UBERON:0002048 |
| | Bronchus | UBERON:0002185 |
| | Sputum | UBERON:0007311 |
| Upper GI tract | Stomach | UBERON:0000945 |
| | Mucosa of stomach | UBERON:0001199 |
| | Duodenum | UBERON:0002114 |
| | Duodenal mucosa | UBERON:0000320 |
| Lower GI tract | Colon | UBERON:0001155 |
| | Colonic mucosa | UBERON:0000317 |
| | Intestine | UBERON:0000160 |
| | Intestinal mucosa | UBERON:0001242 |
| | Large intestine | UBERON:0000059 |
| | Caecum | UBERON:0001153 |
| | Small intestine | UBERON:0002108 |
| | Mucosa of small intestine | UBERON:0001988 |
| | Ileum | UBERON:0002116 |
| | Rectum | UBERON:0001052 |
| | Mucosa of rectum | UBERON:0003346 |
| | Feces | UBERON:0001988 |
| | Meconium | UBERON:0007109 |
| Skin | Skin of body | UBERON:0002097 |
| | Skin of cheek | UBERON:0008803 |
| | Skin of forearm | UBERON:0003403 |
| | Skin of penis | UBERON:0001331 |
| | Skin of sole of pes | UBERON:0013778 |
| | Interdigital space | UBERON:0036252 |

| | | |
|---|---|---|
| Vaginal | Vagina | UBERON:0000996 |
| | Lower part of vagina | UBERON:0015243 |
| | Vaginal fluid | UBERON:0036243 |
| | Posterior fornix of vagina | UBERON:0016486 |
| Female reproductive system | Uterus | UBERON:0000995 |
| | Uterine cervix | UBERON:0000002 |
| | Uterovesical pouch | UBERON:0011049 |
| | Endocervix | UBERON:0000458 |
| | Endometrium | UBERON:0001295 |
| | Ovary | UBERON:0000992 |
| | Placenta | UBERON:0001987 |
| Male reproductive system | Prostate gland secretion | UBERON:0004796 |
| | Semen | UBERON:0001968 |
| Blood | Blood | UBERON:0000178 |
| Breast milk | Milk | UBERON:0001913 |
| Urine | Urine | UBERON:0001088 |
| Lymph node | Mesenteric lymph node | UBERON:0002509 |
| Other | Breast tissue | UBERON:0000310 |
| | Peritoneal fluid | UBERON:0001268 |

**Table S2: Reported measures of alpha diversity.** Shown are the number of experiments that reported one of the indicated alpha diversity measures in the columns with either decreased, increased, or unchanged alpha diversity in the exposed group when compared to the unexposed group.

| | Shannon | Richness | Chao1 | Simpson | Inverse Simpson | Pielou | Total |
|---|---|---|---|---|---|---|---|
| Decreased | 150 | 82 | 98 | 42 | 14 | 11 | 397 |
| Increased | 117 | 93 | 62 | 33 | 6 | 4 | 315 |
| Unchanged | 426 | 235 | 232 | 164 | 30 | 25 | 1122 |
| Total | 703 | 410 | 392 | 239 | 50 | 40 | 1834 |

**Table S3: Body sites with frequently reported changes in alpha diversity.** Shown are the top 5 body sites most frequently reported with increased (top) or decreased (bottom) alpha diversity in the exposed sample group when compared to the unexposed sample group.

| | Increased | Decreased | Unchanged |
|---|---|---|---|
| Saliva | 16 | 6 | 18 |
| Mouth | 11 | 2 | 16 |
| Posterior fornix of vagina | 8 | 0 | 3 |
| Uterine cervix | 9 | 1 | 30 |
| Vagina | 9 | 3 | 24 |
| Feces | 72 | 132 | 485 |
| Stomach | 3 | 15 | 4 |
| Skin of body | 2 | 7 | 14 |
| Caecum | 1 | 4 | 2 |
| Rectum | 0 | 2 | 11 |

**Table S4: Conditions with frequently reported changes in alpha diversity.** Shown are the top 5 conditions most frequently reported with increased (top) or decreased (bottom) alpha diversity in the exposed sample group when compared to the unexposed sample group.

| | Increased | Decreased | Unchanged |
|---|---|---|---|
| Air pollution | 14 | 5 | 9 |
| Human papilloma virus infection | 8 | 1 | 33 |
| Cervical cancer | 5 | 0 | 5 |
| Hypertension | 4 | 0 | 2 |
| Periodontitis | 4 | 0 | 5 |
| COVID-19 | 7 | 28 | 38 |
| Antimicrobial agent | 5 | 20 | 39 |
| Gastric cancer | 2 | 15 | 16 |
| Chronic kidney disease | 0 | 5 | 2 |
| Graft versus host disease | 2 | 7 | 4 |

**Table S5: Individual CRC studies from `curatedMetagenomicData` (cMD) for which signatures of differential abundant taxa are included in BugSigDB.**

| cMD dataset | PMID | Study (BugSigDB) | Comments |
|---|---|---|---|
| ZellerG_2014 | 25432777 | Study 595 | no taxa on genus or species level |
| VogtmannE_2016 | 27171425 | Study 612 | one taxon on genus or species level |
| YachidaS_2019 | 31171880 | Study 630 | 258 CRC vs 251 healthy samples |
| FengQ_2015 | 25758642 | Study 631 | 41 CRC vs 55 healthy samples |

**Table S6: Ranking of the signatures of FengQ_2015 and YachidaS_2019 in an over-representation analysis of 776 BugSigDB signatures.**

| cMD dataset | samples | genera | DA genera | FDR | Rank |
|---|---|---|---|---|---|
| YachidaS_2019 | 509 | 13 | 10 | $2.5 \cdot 10^{-5}$ | 13 |
| FengQ_2015 | 96 | 10 | 4 | 0.16 | 281 |
